# Does Blood-Brain Barrier Disruption Define the Glioma Extracellular Metabolome?

**DOI:** 10.1101/2021.08.24.21262320

**Authors:** Cecile Riviere-cazaux, Lucas P. Carlstrom, Karishma Rajani, Amanda Munoz-casabella, Masum Rahman, Ali Gharibi-Loron, Desmond A. Brown, Kai J Miller, Jaclyn J. White, Benjamin T. Himes, Ignacio Jusue-Torres, Samar Ikram, Seth Ransom, Renee Hirte, Ju-Hee Oh, William F. Elmquist, Jann N. Sarkaria, Rachael A. Vaubel, Moses Rodriguez, Arthur Warrington, Sani H. Kizilbash, Terry C. Burns

## Abstract

**Background:** The extracellular microenvironment modulates cancer behavior. Although radiographic contrast enhancement is an ominous finding in gliomas, it remains unclear if the associated blood-brain barrier disruption merely reflects or functionally supports tumor aggressiveness.

**Methods:** We utilized intra-operative microdialysis to sample the extracellular metabolome of radiographically diverse regions during fifteen neurosurgical resections. The global extracellular metabolome of recovered microdialysate was evaluated via ultra-performance liquid chromatography tandem mass spectrometry (UPLC-MS/MS) and assessed using enrichment and correlation analyses.

**Results:** Among 162 named metabolites identified via ultra-performance liquid chromatography tandem mass spectrometry, guanidinoacetate (GAA), was 126.32x higher in enhancing tumor than in adjacent brain. 48 additional metabolites were 2.05-10.18x more abundant in enhancing tumor than brain. With exception of GAA, and 2-HG in IDH-mutant gliomas, differences between non-enhancing tumor and brain microdialysate were comparatively modest and less consistent. The enhancing but not the non-enhancing glioma metabolome was significantly enriched for plasma-associated metabolites largely comprising amino acids and carnitines.

**Conclusions:** Our findings suggest that metabolite diffusion through a disrupted blood-brain barrier may largely define the enhancing extracellular glioma metabolome. Future studies are needed to determine how the altered extracellular metabolome impacts glioma behavior.

## Introduction

Astrocytomas and oligodendrogliomas comprise most adult primary malignant brain tumors and remain incurable, regardless of the best available therapies^1^. Isocitrate dehydrogenase (IDH) mutations occur in all oligodendrogliomas and a subset of astrocytomas, affording improved prognosis due to typically slower growth and more favorable response to standard-of-care radiation and alkylating chemotherapy^2,3^. Glioma heterogeneity hampers therapeutic generalizations across diverse patient cohorts. Moreover, diverse genetic phenotypes accumulate within individual patients’ tumor ecosystems, hampering therapeutic efforts to target glioma-associated mutations^4,5^. Conversely, molecularly diverse gliomas may resort to similar metabolic pathways in the setting of nutrient deprivation, hypoxia, and genotoxic stress^6,7^. Although glioma metabolism is increasingly scrutinized for therapeutic targets, few strategies currently exist to interrogate the metabolic microenvironment of human gliomas *in situ*.

Microdialysis has been used to quantify human extracellular biomarkers of traumatic and hypoxic brain injury in neurocritical care units. Microdialysis is also a well-established method to quantify central nervous system (CNS) drug delivery in early phase clinical trials^8-10^ and has been used longitudinally to study metabolites present in human gliomas when compared to adjacent brain^11,12^. However, while some studies have utilized intraoperative low molecular weight microdialysis (8-20 kDA) to quantify select extracellular metabolites^13-15^, no study thus far has deployed intra-operative microdialysis to characterize the global extracellular glioma metabolome, nor to compare the metabolome of contrast-enhancing versus non-enhancing tumor regions. We used variable flow-rate pumps set to 2 µL/min and high molecular weight catheters (100 kDa) to maximize the volume of microdialysate recovered during 15 surgeries in 14 patients. We report that contrast-enhancing (radiographically blood-brain barrier (BBB) disrupted) regions of high-grade gliomas (HGGs) exhibited a conserved extracellular metabolome across both IDH-mutant and IDH-WT astrocytomas, similar to findings from other groups utilizing intra-or-post-operative microdialysis^14,16,17^. However, comparison to the radiographically BBB intact (non-enhancing) portions of these tumors revealed that this enhancing glioma signature was significantly enriched for plasma-associated metabolites, suggesting that BBB disruption may contribute to the glioma extracellular metabolome.

## MATERIALS AND METHODS

### Patient Cohort, Study Design, and Intraoperative Microdialysis

All study procedures were approved by the Mayo Clinic Institutional Review Board (IRB). Patients provided written informed consent to participate in NCT04047264--an ongoing study utilizing intraoperative high molecular weight (100 kDa) microdialysis under an investigational device exemption. Study eligibility included adults (>18yo) undergoing a clinically indicated surgery for known or suspected glioma. Each patient (n=14; 15 surgeries; **Table 1**) underwent intraoperative microdialysis using up to three 100 kDA catheters and variable rate microdialysis pumps (M Dialysis 71 High Cut-Off Brain Microdialysis Catheters and 107 Microdialysis Pump, respectively) across radiographically diverse regions (**Fig. 1A-B**). The target and trajectory of each microdialysis catheter was planned prior to surgery using the computerized neuronavigation system. When feasible, the microdialysis membrane (typically 10 mm) of each catheter was planned to be located fully within (i) enhancing tumor (typically “catheter X”), (ii) non-enhancing tumor (typically “catheter Y”), or (iii) adjacent brain planned for inclusion within the resection volume (typically “catheter Z”). Once the dura was opened and the cortical surface exposed, pia at each planned entry point was coagulated and opened with microscissors. The pre-flushed catheter was then advanced to the intended trajectory. Microdialysis was performed at 2 µL/min based on a prior intra-operative study demonstrating that this flowrate could enable sufficient collection of analytes for mass spectrometry^15^, with collection vials exchanged every 20 minutes. See supplemental methods for details of microdialysis procedure, pathology, and molecular tumor analyses. Each patient is referred to via a unique de-identified label reflecting their histology, IDH-status, and WHO grade according to 2021 WHO classification^18^ (Table 1).

**Table 1.** Intraoperative microdialysis patient characteristics. The patient IDs and final pathologic diagnoses, including IDH-status and other available key molecular features, are summarized together with the catheter locations. Additional specifics regarding each patient, tumor, and catheter are available in Supplemental Table 1. *Diagnosis of IDH-WT GBM was reached for patient GBM^WT^2/2B without need for further molecular studies. †: Region of minimal enhancement, suggestive of tumor necrosis.

| Patient ID | Pathology | IDH Status | Other molecular/genetic features | Catheter X | Catheter Y | Catheter Z |
| --- | --- | --- | --- | --- | --- | --- |
| Oligo<sup>2</sup> | Oligodendroglioma WHO 2 | Mutant | 1p/19q co-deletion; TERT-mutant | Non-enhancing (NE) | Non-enhancing (NE) | Non-enhancing (NE) |
| Oligo<sup>3</sup>1 | Oligodendroglioma WHO 3 | Mutant | 1p/19q co-deletion; TERT-mutant | Non-enhancing (NE) | Non-enhancing (NE) | Brain adjacent to tumor (B) |
| Oligo<sup>3</sup>2 | Oligodendroglioma WHO 3 | Mutant | 1p/19q co-deletion; TERT-mutant | Non-enhancing (NE) | Non-enhancing (NE) | Brain adjacent to tumor (B) |
| Oligo<sup>3</sup>3 | Oligodendroglioma WHO 3 | Mutant | 1p/19q co-deletion; TERT-mutant | Non-enhancing (NE) | Non-enhancing (NE) | Brain adjacent to tumor (B) |
| GemAstro<sup>3-mut</sup> | Gemistocytic Astrocytoma WHO 3 | Mutant | None remarkable | Non-enhancing (NE) | Non-enhancing (NE) | Brain adjacent to tumor (B) |
| Astro<sup>4-mut</sup>1 | Astrocytoma WHO 4 (Recurrent) | Mutant | MGMT non-methylated; loss of CDKN2A/B; complex karyotype | Enhancing (E) | Non-enhancing (NE) | Brain adjacent to tumor (B) |
| Astro<sup>4-mut</sup>2 | Astrocytoma WHO 4 | Mutant | MGMT non-methylated; loss of CDKN2A/B | Enhancing (E) | Non-enhancing (NE) | Brain adjacent to tumor (B) |
| Astro<sup>4-mut</sup>3 | Astrocytoma WHO 4 (Recurrent) | Mutant | Indeterminate MGMT methylation | Enhancing (E) | Enhancing† (E<sup>†</sup>) | Brain adjacent to tumor (B) |
| H3K27M-Astro<sup>4-WT</sup> | Infiltrating Astrocytoma WHO 4 | Wild-type | H3K27M, TERT-mutant; MGMT non-methylated | Enhancing (E) | Non-enhancing (NE) | Brain adjacent to tumor (B) |
| GBM<sup>WT</sup>1 | Glioblastoma | Wild-type | MGMT non-methylated; loss of CDKN2A/B; TERT-mutant | Enhancing (E) | Non-enhancing (NE) | Brain adjacent to tumor (B) |
| GBM<sup>WT</sup>2 | Glioblastoma (Primary) | Wild-type | Indeterminate MGMT methylation* | Enhancing (E) | Non-enhancing (NE) | Brain adjacent to tumor (B) |
| GBM<sup>WT</sup>2B | Glioblastoma (Recurrent) | Wild-type | Indeterminate MGMT methylation* | Enhancing (E) |  | Brain adjacent to tumor (B) |
| GBM<sup>WT</sup>3 | Glioblastoma | Wild-type | MGMT non-methylated | Enhancing (E) | Non-enhancing (NE) | Brain adjacent to tumor (B) |
| GBM<sup>WT</sup>4 | Glioblastoma (Recurrent) | Wild-type | MGMT non-methylated; EGFR amplification; loss of CDKN2A/B | Enhancing (E) | Non-enhancing (NE) | Brain adjacent to tumor (B) |
| MolecGBM<sup>WT</sup> | Molecular Glioblastoma | Wild-type | Gain of ch. 7, loss of ch. 10; TERT-mutant; MGMT non-methylated | Non-enhancing (NE) | Non-enhancing (NE) | Brain adjacent to tumor (B) |

**Figure 1.**
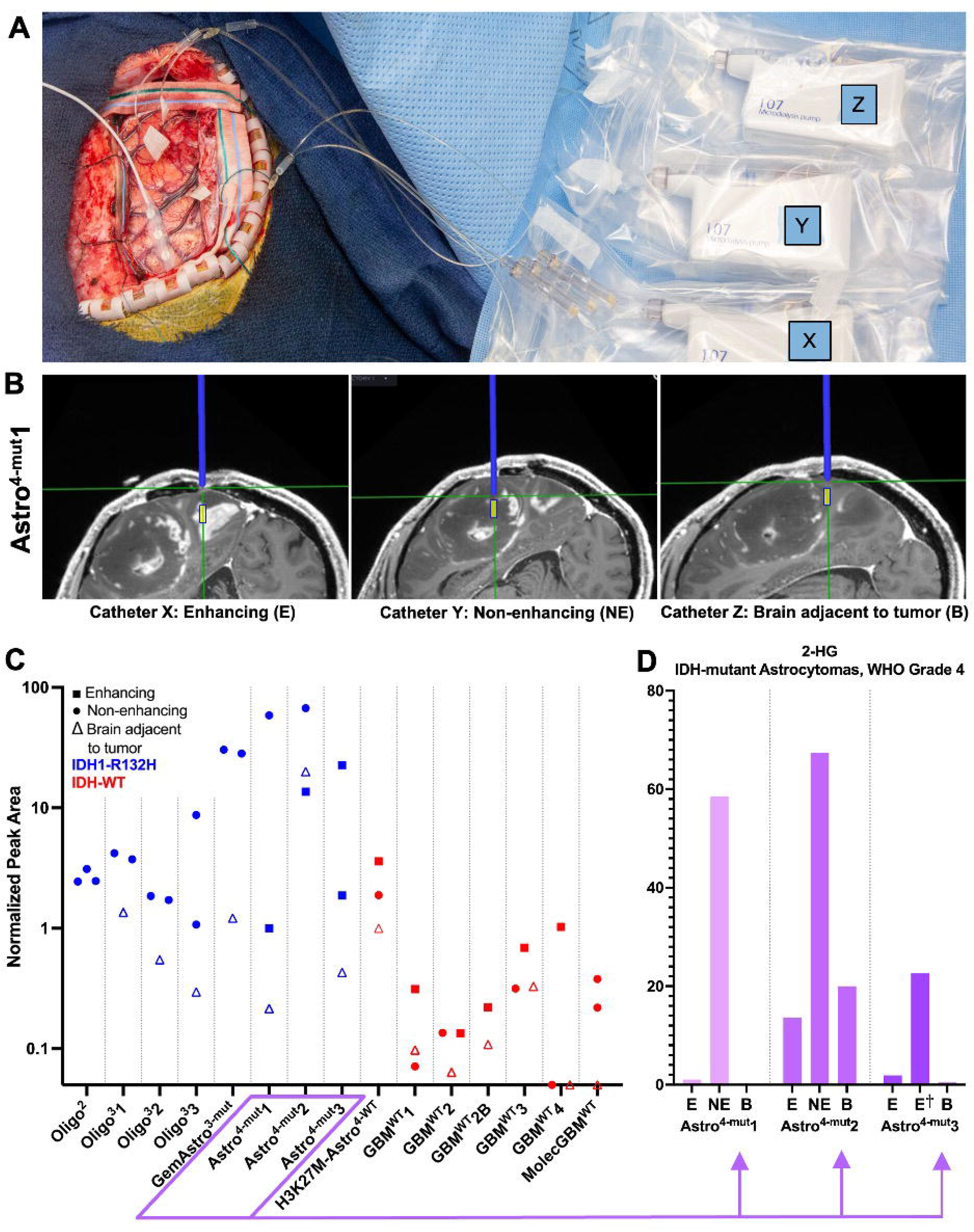
Intraoperative microdialysis set-up and D-2-hydroxyglutarate (D-2-HG) detection. **(A)** A representative intraoperative photograph (Oligo^2^) demonstrating the experimental setup, including placement of three 100 kDA microdialysis catheters, as well as pumps (X, Y, Z) and collection vials within the surgical field. **(B)** Illustrative intraoperative trajectory views captured from the Neuronavigation system (Astro^4-mut^1) demonstrate the planned trajectory (blue line) toward the intended target location for each 10 mm microdialysis membrane (indicated by yellow box) in enhancing (E, Catheter X) and non-enhancing tumor (NE, Catheter Y), in addition to relatively normal brain adjacent to tumor (B, Catheter Z). **(C)** 2-HG peak areas from UPLC-MS/MS were measured in the microdialysate from each of the 44 catheters. IDH status is indicated as mutant (blue) or wild type (red). Symbol shape indicates catheter placement location (see legend). **(D)** The 2-HG peak areas from three patients with enhancing grade 4 IDH-mutant astrocytomas are shown in closer detail.

**Figure 2.**
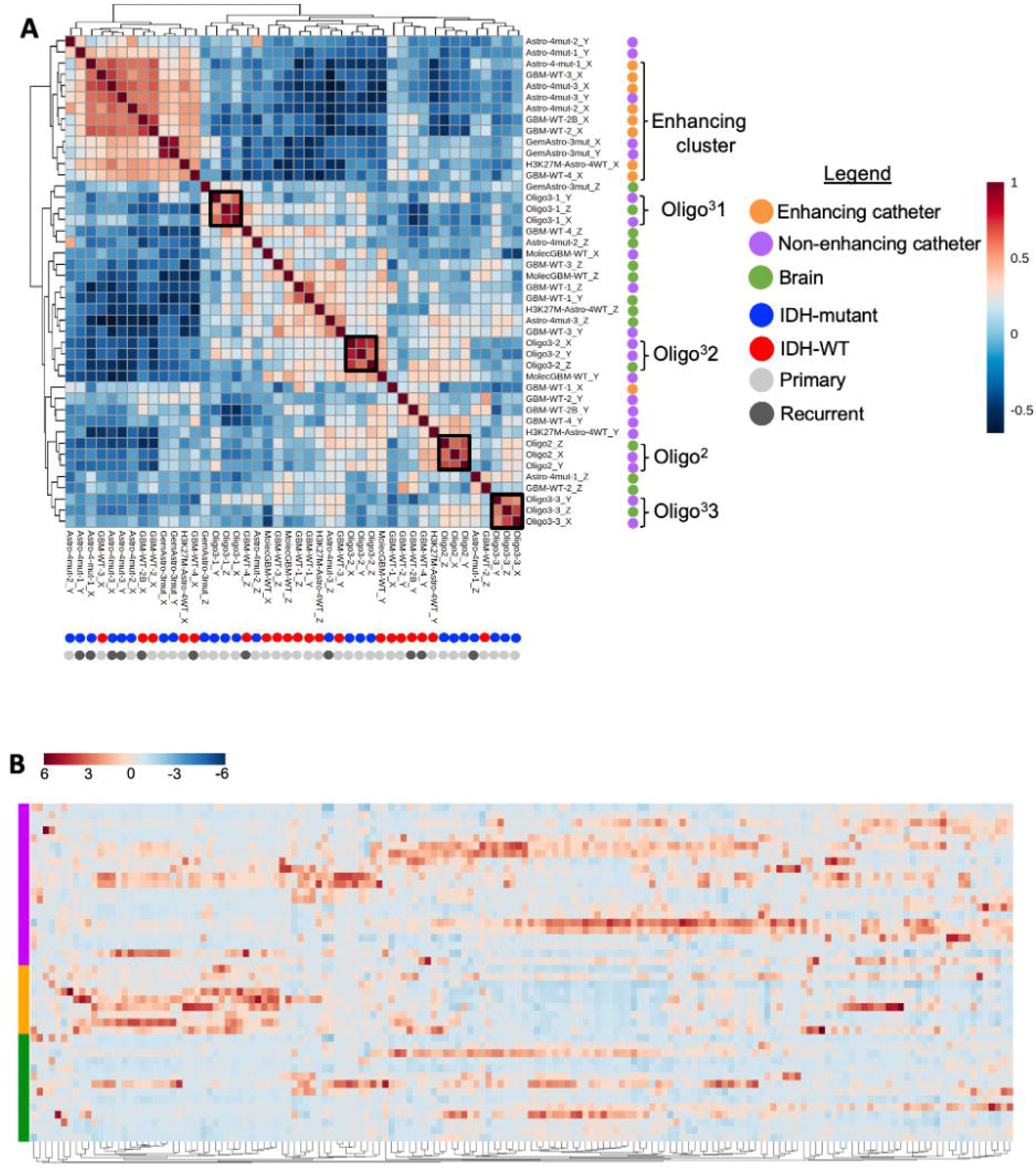
Microdialysate samples cluster based on patient identity and catheter location. **(A)** Spearman correlation map of the 44 catheters from all 15 microdialysis cases (14 patients). Dot colors indicate catheter location, IDH status, and primary or recurrent status. (Minimal correlation: -0.7, maximal correlation: 1). **(B)** Hierarchical clustering heat map (Ward method, Euclidean distance measure) of the 44 catheters based on 162 metabolites present in at least 90% (40/44) catheters. (Normalized abundance by MetaboAnalyst: minimum, -6 to maximum, 6).

### Targeted Analysis of D/L-2-HG

Targeted metabolomic analysis of microdialysate was performed by the Mayo Clinic metabolomic core facility. L and D isomers of 2-hydroxyglutaratic acid were derivatized and quantified by liquid chromatography mass spectrometry (LC/MS) using slight modifications to previously described methods^19-21^. See supplemental methods for details.

### Untargeted Metabolomic Analysis

Untargeted metabolomic analysis was performed by Metabolon, Inc. Ultra-performance liquid chromatography tandem mass spectrometry (UPLC-MS/MS), which combines physical separation of liquid chromatography with the mass analysis capabilities of mass spectrometry. See supplemental methods for details.

### Ranked Metabolite lists

Ranked metabolite lists of tumor (Catheter X or Y) versus brain (Catheter Z) were generated from fifteen cases based on calculated fold changes of the peak area (e.g. X/Z or Y/Z) for each metabolite. A tumor versus brain ranked metabolite list could not be generated for one patient (Oligo^2^) given all catheters were located within tumor. In patients with both enhancing and non-enhancing tumor catheters (n=7 patients), a ranked metabolite lists of enhancing vs. non-enhancing tumor was generated (Catheter X/Y). Initial analysis was performed using samples generated during the first 5 surgeries. Three of these patients had contrast-enhancing tumors. To determine an overall enhancing tumor (E)-to-brain (B) ranking for each metabolite, each metabolite present in both catheters was ranked based on fold change (E/B) and then averaged across all three patients for that metabolite. In the remaining cases, the individual tumor-to-brain rankings were compared against the first cohort’s average enhancing tumor-to-brain ranking of metabolites. Similar analyses were performed for enhancing versus non-enhancing tumor (E/NE) and non-enhancing tumor versus brain (NE/B). To understand the potential impact of extracellular blood-derived metabolites on microdialysate metabolic signatures, we additionally generated a ranked list of bloodiness-associated metabolites using a pair of cranial CSF samples obtained from a single patient immediately after incising dura, prior to resection of a 4^th^ ventricular ependymoma. Both samples were obtained within approximately 1 minute of each other—one with and one without blood contamination from the surgical wound. Samples were centrifuged at 400 Gy for 10 minutes to remove red blood cells and other debris, and aliquots of supernatant submitted to metabolon for global metabolomic analysis. The ranked metabolite list was based on metabolite fold-change (bloody / non-bloody) between samples, after filtering for metabolites present in both CSF and >90% of microdialysate samples.

### Enrichment Analysis

Gene Set Enrichment Analysis (GSEA) is the most widely utilized analytical method for rank-based analysis of data sets in biomedicine. GSEA is frequently performed using curated gene sets from the molecular signatures database (MSigDB), though custom analyte sets can be used enabling enrichment analysis of metabolites rather than genes. In GSEA, custom analyte sets are queried against a ranked list of analytes to determine where those analytes fall on the ranked list (**Supplementary Fig. 1**). If the analytes are most frequently found at the top of the ranked list, this is known as “positive enrichment;” if they fall at the end of the ranked list, this is known as “negative enrichment.” The software uses this information to calculate a normalized enrichment score and a p value. FDR values are also provided to correct for multiple hypothesis testing when multiple analyte sets are evaluated simultaneously for enrichment in a particular data set. Custom metabolite libraries were created using the top and bottom 35 metabolites from each ranked metabolite list described above and can be found in **Supplementary Data** (.gmx file). Using these metabolite libraries (.gmx file), the full ranked list for each patient (Catheter X vs. Z, Catheter Y vs. Z, or Catheter X vs Y .rnk files) was then used for enrichment analysis to determine each patient’s relative positive and negative enrichment for other patients’ tumor or brain extracellular metabolomes based on the metabolites sets in the .gmx file (**Supplemental Data**). Additionally, results from Björkblom et al. were included as a metabolite set to provide an external reference^17^. The bloody and non-bloody CSF metabolite libraries were included to identify the relative enrichment of tumor or brain microdialysate for plasma-derived metabolites.

### MetaboAnalyst

Metabolomic analyses were performed using MetaboAnalyst (5.0), a publicly available web-based tool to analyze and visualize metabolomic data^22^. The normalized peak area data for the metabolites present across at least 40/44 catheters were entered into MetaboAnalyst without further filtering. Assuming a monotonical rather than linear relation between metabolites and samples, Spearman Correlation maps were generated in MetaboAnalyst based on features (metabolites present across all samples) and samples.

### Statistical Analysis

Data are presented as normalized peak areas (median = 1 for each metabolite within each analyzed batch. Raw and normalized peak area data are provided in Supplemental File 1. MetaboAnalyst 5.0 was used for Spearman correlations. Enrichment Analysis was performed using GSEA 4.1.0 (Gene Set Enrichment Analysis), repurposed for metabolite set analysis using custom metabolite sets. The normal distribution of metabolites in Figure 4 and Table 2 was tested via D’Agostino-Pearson test; a Wilcoxon signed rank test for non-parametric distributions was then performed on paired enhancing tumor and brain metabolites using GraphPad PRISM 9.1. Volcano plot cut-offs were set at FC≥2 and p≤0.05. Graphs were generated using GraphPad PRISM 9.1. FDR≤0.05 was considered statistically significant for enrichment analysis.

**Table 2.** Significantly altered metabolites across enhancing, non-enhancing, and brain microenvironments in paired patient specimens (n=9 for E/B and n=7 for E/NE or NE/B). *bold = p<0.05.

| Metabolite | FC (E/B) | P-value | FC (E/NE) | P-value | FC (NE/B) | P-value |
| --- | --- | --- | --- | --- | --- | --- |
| Guanidinoacetate (GAA) | **126.32** | 0.0039 | 9.76 | 0.2188 | **13.63** | 0.0469 |
| proline | **10.18** | 0.0039 | **4.46** | 0.0313 | 2.09 | 0.1094 |
| deoxycarnitine | **8.71** | 0.0039 | **4.55** | 0.0156 | 1.94 | 0.3750 |
| carnitine | **7.65** | 0.0039 | **3.32** | 0.0313 | 2.18 | 0.0781 |
| acetylcarnitine (C2) | **7.47** | 0.0039 | 2.76 | 0.1094 | **2.97** | 0.0469 |
| N1-methyl-2-pyridone-5-carboxamide (2-PY) | **7.18** | 0.0391 | **3.20** | 0.0156 | 1.28 | 0.6875 |
| N6-methyllysine | **7.17** | 0.0039 | **4.67** | 0.0313 | 1.16 | 0.2969 |
| methionine | **7.09** | 0.0039 | **2.94** | 0.0156 | 2.41 | 0.0781 |
| dimethylglycine | **6.90** | 0.0039 | **3.75** | 0.0156 | 1.51 | 0.4688 |
| asparagine | **6.84** | 0.0039 | **3.64** | 0.0156 | 1.55 | 0.3750 |
| propionylcarnitine (C3) | **6.78** | 0.0078 | **3.77** | 0.0469 | 1.84 | 0.1094 |
| alanine | **6.54** | 0.0039 | **3.11** | 0.0156 | 1.96 | 0.0781 |
| valine | **6.01** | 0.0039 | **3.24** | 0.0156 | 1.70 | 0.1563 |
| tyrosine | **5.66** | 0.0039 | **3.14** | 0.0313 | 1.62 | 0.2969 |
| glycine | **5.66** | 0.0078 | 2.43 | 0.2188 | 2.18 | 0.1094 |
| imidazole lactate | **5.57** | 0.0039 | **3.78** | 0.0156 | 1.09 | 0.5781 |
| urate | **5.05** | 0.0039 | **2.63** | 0.0156 | 1.60 | 0.0781 |
| lysine | **4.54** | 0.0039 | **2.68** | 0.0156 | 1.41 | 0.2969 |
| allantoin | **4.52** | 0.0039 | **3.96** | 0.0313 | 0.90 | 0.5781 |
| histidine | **4.49** | 0.0039 | **2.30** | 0.0313 | **1.73** | 0.0156 |
| phenylalanine | **4.48** | 0.0039 | **2.59** | 0.0156 | 1.52 | 0.0781 |
| trigonelline (N'-methylnicotinate) | **4.40** | 0.0078 | 1.75 | 0.1563 | 2.57 | 0.2969 |
| 3-hydroxybutyrate (BHBA) | **4.40** | 0.0039 | **2.22** | 0.0313 | **1.77** | 0.0313 |
| 3-methyl-2-oxobutyrate | **4.34** | 0.0039 | 3.86 | 0.0781 | 0.80 | 0.5781 |
| hydroxyproline | **4.22** | 0.0039 | **2.11** | 0.0313 | 1.48 | 0.2188 |
| threonine | **4.20** | 0.0039 | **2.27** | 0.0313 | 1.58 | 0.1563 |
| leucine | **4.12** | 0.0039 | **2.43** | 0.0156 | 1.54 | 0.0781 |
| tryptophan | **3.96** | 0.0039 | **2.33** | 0.0313 | 1.40 | 0.2969 |
| N1-methyl-4-pyridone-3-carboxamide (4-PY) | 3.88 | 0.0742 | **2.16** | 0.0156 | 1.27 | 0.5781 |
| N,N,N-trimethyl-5-aminovalerate | **3.88** | 0.0039 | 2.17 | 0.0781 | 1.74 | 0.0781 |
| isoleucine | **3.76** | 0.0039 | **2.29** | 0.0156 | 1.52 | 0.0781 |
| guanine | **3.73** | 0.0078 | 1.24 | 0.5781 | **2.62** | 0.0469 |
| N,N,N-trimethyl-alanylproline betaine (TMAP) | **3.50** | 0.0039 | 1.59 | 0.1094 | 1.58 | 0.2969 |
| p-cresol sulfate | **3.36** | 0.0039 | 2.03 | 0.2969 | 1.44 | 0.2188 |
| Glutamine degradant | **3.24** | 0.0078 | **1.88** | 0.0156 | 1.61 | 0.0781 |
| arginine | **3.16** | 0.0039 | **2.17** | 0.0156 | 1.25 | 0.6875 |
| Lactate* | **3.12** | 0.0313 | 1.34 | 0.0625 | 2.32 | 0.0625 |
| ornithine | **3.07** | 0.0039 | **2.36** | 0.0313 | 1.13 | 1.0000 |
| taurine | **2.99** | 0.0117 | 1.76 | 0.1094 | **2.08** | 0.0156 |
| thioprolin | **2.97** | 0.0078 | 1.31 | 0.0781 | 2.50 | 0.0781 |
| gamma-glutamylglutamine | **2.93** | 0.0117 | **1.45** | 0.0469 | **2.03** | 0.0313 |
| serine | **2.80** | 0.0039 | **2.12** | 0.0469 | 1.12 | 0.9375 |
| N6,N6,N6-trimethyllysine | **2.62** | 0.0195 | 1.80 | 0.0781 | 1.27 | 0.2188 |
| glycerophosphorylcholine (GPC) | 2.60 | 0.0742 | 0.87 | 1.0000 | **4.32** | 0.0156 |
| phenol sulfate | **2.59** | 0.0078 | 2.08 | 0.0781 | 1.09 | 0.2188 |
| N-acetylputrescine | **2.44** | 0.0273 | 1.30 | 0.8125 | 1.75 | 0.4688 |
| ergothioneine | **2.41** | 0.0391 | 0.60 | 0.3750 | **3.26** | 0.0156 |
| 3-hydroxyisobutyrate | **2.27** | 0.0391 | 1.51 | 0.3750 | 1.15 | 0.3750 |
| stachydrin | **2.24** | 0.0078 | 1.45 | 0.2188 | 1.39 | 0.0781 |
| 1-methyl-5-imidazolelactate | 2.13 | 0.1073 | 0.56 | 0.8125 | **4.27** | 0.0156 |
| 7-methylguanine | **2.10** | 0.0195 | 1.53 | 0.3750 | 1.40 | 0.2945 |
| N-acetyl glycine | **2.05** | 0.0039 | 1.71 | 0.1563 | 1.10 | 0.2188 |
| glycerophosphoglycerol | 2.04 | 0.2340 | 0.45 | 0.4688 | **5.21** | 0.0156 |
| malate | 1.87 | 0.1641 | 0.70 | 0.9375 | **2.99** | 0.0156 |
| phosphoethanolamine (PE) | 1.56 | 0.6523 | 0.49 | 0.5781 | **4.83** | 0.0156 |
| creatine | 1.05 | 0.5703 | 0.55 | 0.2188 | **2.25** | 0.0313 |
| glycerol-3-phosphate | 1.02 | 0.5703 | 0.40 | 0.3750 | **3.25** | 0.0313 |
| phosphate | 0.87 | 1.0000 | **0.33** | 0.0313 | **2.91** | 0.0469 |
| glycerophosphoethanolamine | 0.82 | 0.8203 | 0.43 | 0.1563 | **2.83** | 0.0156 |
| inosine | 0.64 | 0.4258 | **0.48** | 0.0156 | 1.60 | 0.1563 |
| myo-inositol | 0.51 | 0.4258 | **0.46** | 0.0469 | 1.13 | 0.5781 |
| orotate | 0.47 | 0.1641 | **0.39** | 0.0313 | 0.98 | 0.8125 |
| N-formylmethionine | 0.42 | 0.1289 | **0.36** | 0.0469 | 1.13 | 0.6875 |
| aspartate | **0.40** | 0.0195 | 0.43 | 0.2188 | 1.06 | 0.6875 |
| lyxonate | **0.39** | 0.0078 | 0.47 | 0.0781 | 0.86 | 0.9375 |
| N-acetyl asparagine | **0.37** | 0.0039 | **0.37** | 0.0469 | 0.86 | 0.6875 |
| 3-hydroxy-3-methylglutarate | **0.37** | 0.0391 | 0.29 | 0.0781 | 1.55 | 0.1563 |
| N-acetylglutamine | **0.34** | 0.0078 | **0.34** | 0.0156 | 0.99 | 0.6875 |
| N-acetylthreonine | **0.34** | 0.0078 | **0.34** | 0.0469 | 0.93 | 1.0000 |
| N-acetyl-beta-alanine | **0.30** | 0.0391 | 0.45 | 0.1422 | 0.73 | 0.6875 |
| N-acetylalanine | **0.30** | 0.0391 | **0.25** | 0.0156 | 0.96 | 0.9375 |
| 2-methylcitrate/homocitrate | **0.29** | 0.0039 | **0.27** | 0.0156 | 0.94 | 0.6875 |
| N-acetylneuraminate | **0.25** | 0.0039 | 0.32 | 0.0781 | 0.75 | 0.9375 |
| fructose | **0.25** | 0.0039 | **0.40** | 0.0469 | 0.56 | 0.4688 |
| N1-methylinosine | **0.23** | 0.0078 | **0.20** | 0.0313 | 1.09 | 0.2969 |
| N-acetylserine | **0.21** | 0.0039 | **0.23** | 0.0313 | 0.87 | 0.6875 |
| Erythronate | **0.18** | 0.0039 | **0.21** | 0.0156 | 0.83 | 1.0000 |
| N-acetylaspartate (NAA) | **0.17** | 0.0039 | **0.16** | 0.0156 | 1.18 | 0.5781 |
| N-acetyl-aspartyl-glutamate (NAAG) | **0.16** | 0.0078 | 0.19 | 0.1563 | 0.73 | 0.6875 |
| 4-acetamidobutanoate | **0.15** | 0.0273 | 0.28 | 0.1563 | 0.96 | 0.9375 |
| 2,3-dihydroxy-5-methylthio-4-pentenoate (DMTPA) | **0.15** | 0.0039 | **0.24** | 0.0156 | 0.55 | 0.0781 |
| 2-O-methylascorbic acid (2-O-MA) | **0.14** | 0.0039 | **0.15** | 0.0156 | 0.83 | 0.8125 |
| ribonate | **0.11** | 0.0039 | **0.13** | 0.0156 | 0.82 | 0.9375 |
| arabonate/xylonate | **0.08** | 0.0039 | **0.14** | 0.0156 | 0.70 | 0.4688 |
| arabitol/xylitol | **0.04** | 0.0039 | **0.08** | 0.0156 | 0.57 | 0.3750 |

\*Fold-change and p-value for lactate are based on n=7 cases, rather than n=9, as the perfusate solution contained lactate in 2 cases.

**Figure 3.**
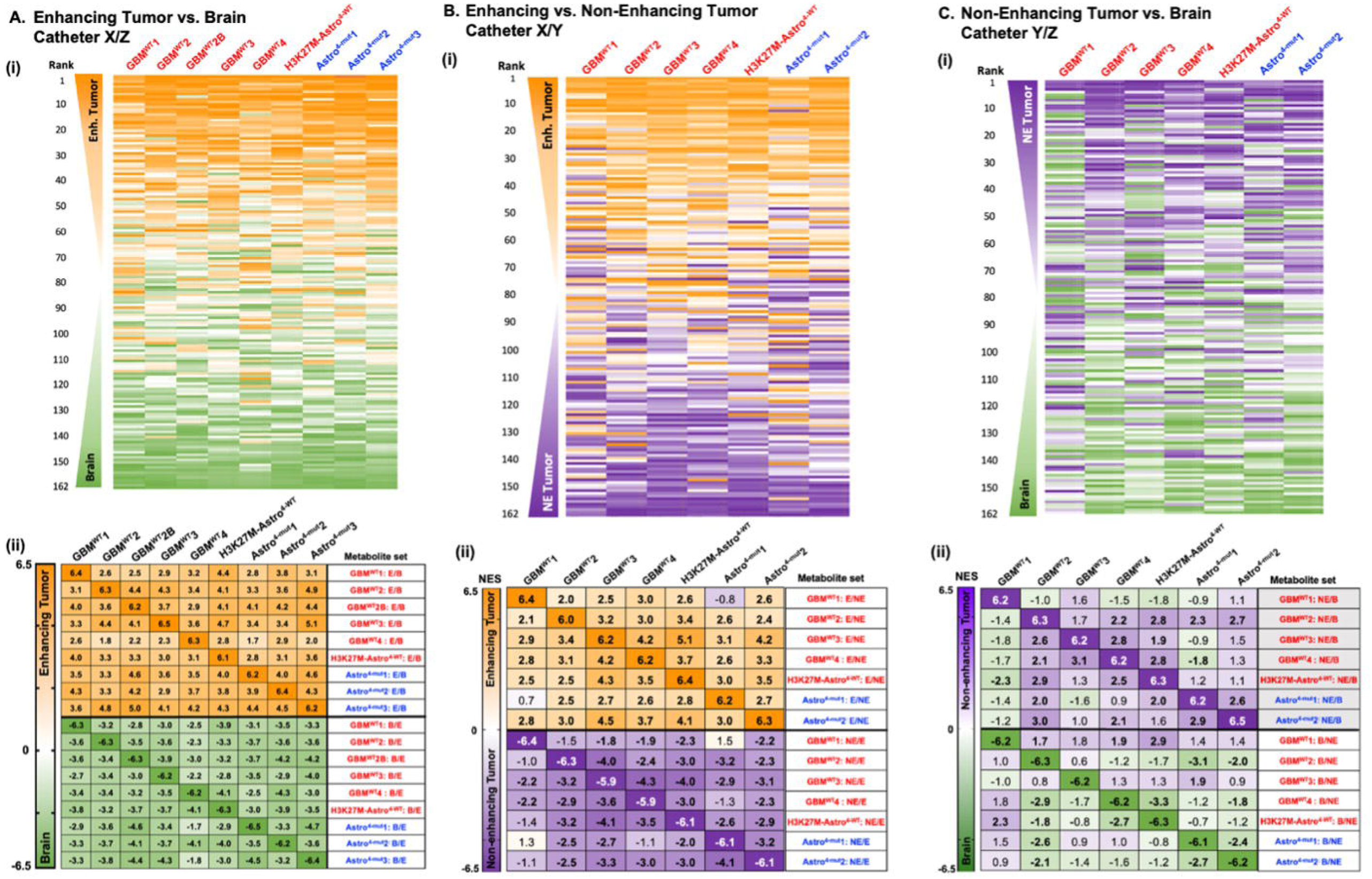
Metabolic signatures of enhancing versus non-enhancing tumors versus brain. **(A)** (i) 162 metabolites present in at least 40/44 catheters were ranked according to the enhancing tumor-versus-brain fold change in each patient. The rank order of each metabolite in each 2-catheter tumor/brain comparison (e.g., catheter X versus Z) is conveyed as a heat map from 1 (orange, enhancing tumor) to 162 (green, brain). Metabolites are listed based on the average of ranks of enhancing vs. brain across nine cases. (ii) Normalized enrichment scores (NES) are shown based upon enrichment analysis for the top 35 enhancing tumor (E) and brain (B)-associated metabolites in the ranked metabolite of enhancing tumor vs. brain (Cath. X/Z) (bold: significant at FDR<0.05; Complete FDR details available in Supplemental Table 2). See Supplemental Figure 1 for graphical depiction of how enrichment analyses are performed by repurposing the Gene Set Enrichment Analysis software. **(B)** Using the same method as in 3A(i), the rank order of each metabolite in enhancing-versus-non-enhancing catheter metabolite lists (Catheter X and Y) is depicted as a heat map in the 7 patients for whom both enhancing and non-enhancing catheters were present. Metabolites are listed based on the average of ranks of enhancing versus non-enhancing tumor across seven patients. (ii) As in 3A(ii), using enhancing versus non-enhancing tumor metabolite ranked lists and metabolite sets. NES are shown for the enrichment of each E vs. NE ranked list for other E vs. NE metabolite sets (bold: FDR<0.05; Complete FDR details available in Supplemental table 3). **(C)** (i) As in 3A-B, the ranked order of metabolites in non-enhancing versus brain metabolite lists is shown as a heat map. Metabolites are ranked based on the average rank of NE tumor to brain across 7 patients. (ii) As in prior enrichment analyses, with non-enhancing versus brain ranked lists and metabolite sets and their respective NESs across patients (bold: FDR<0.05; Complete FDR details available in Supplemental table 4).

**Figure 4.**
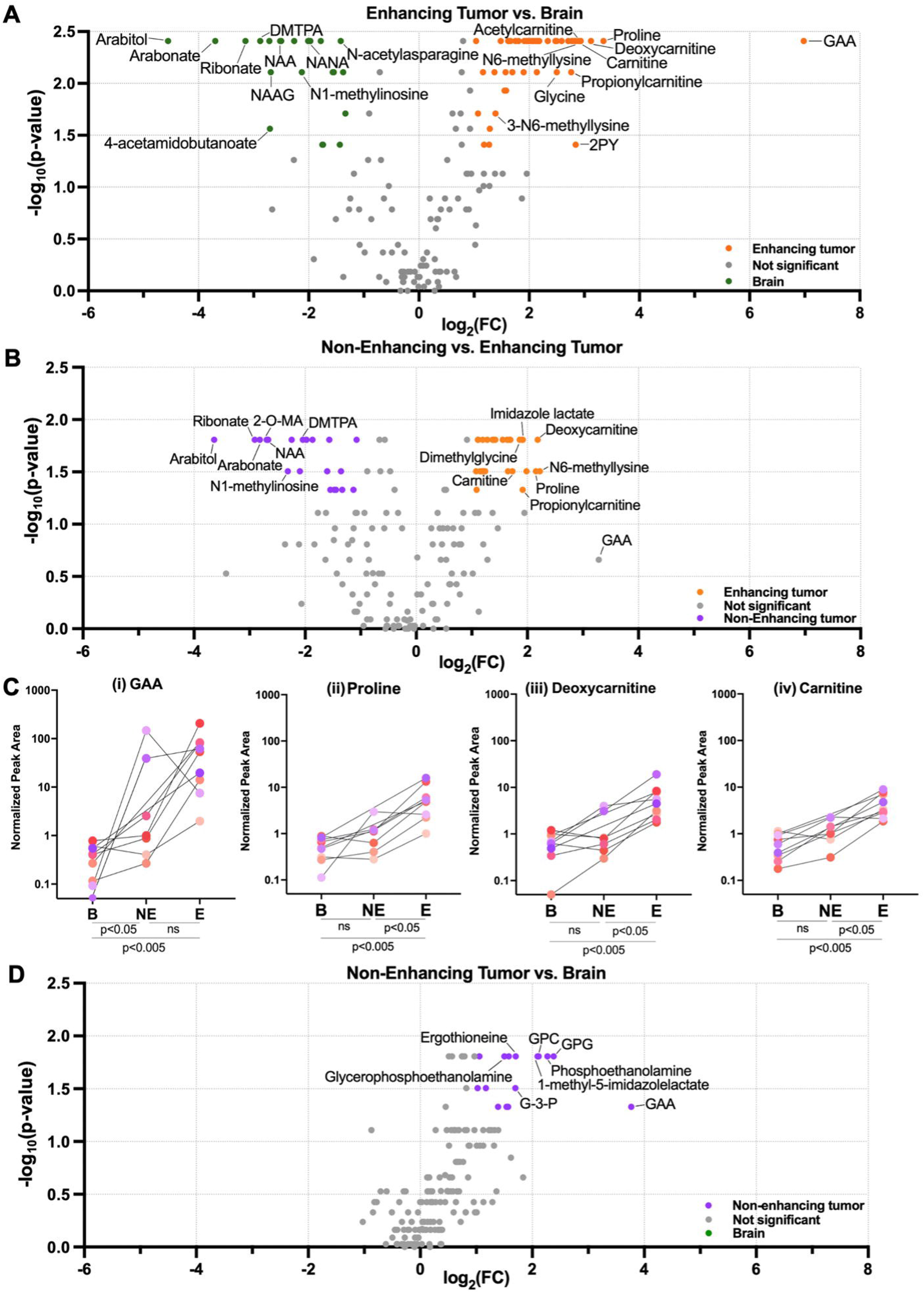
Differentially abundant extracellular metabolites in the enhancing glioma, non-enhancing glioma, and adjacent brain comparisons. Wilcoxon signed-rank tests and fold-changes were calculated between (A) enhancing tumor vs. brain (n=9 paired catheters), (B) enhancing versus non-enhancing tumor (n=7 paired catheters), and (D) non-enhancing tumor versus brain (n=7 paired catheters) were utilized to construct a volcano plot (cut-offs for significance: p-value≤0.05; FC≥2). The normalized peak areas for the top four most differentially abundant extracellular metabolites in enhancing tumor vs. brain are shown in (**C)** for the 9 cases, including the peak area in non-enhancing tumor for the seven patients for whom this catheter was available; colors correlate with patient identity from Table 1. 2-O-MA= 2-O-methylascorbic acid; DMTPA = 2,3-dihydroxy-5-methylthio-4-pentenoic acid; G-3-P=glycerol-3-phosphate; GAA = guanidinoacetate; GPC = glycerophosphocholine; GPG = glycerophosphorylglycerol; NAA = n-acetylaspartate; NAAG = n-acetylaspartylglutamate; NANA = n-acetylneuraminate.

## RESULTS

### Elevated D-2-Hydroxyglutarate (D-2-HG) in IDH-mutant tumor microdialysate

Microdialysis is a method that can be used to sample “dialyzable” extracellular analytes, such as metabolites and certain drugs, from the tumor or brain interstitial space. This is most commonly performed in the post-operative setting with low-molecular weight catheters, including an FDA-approved system consisting of 20 kDA catheters perfused at 0.3 µL/min using the M-dialysis 106 pump. While a limited subset of patients may be willing to undergo post-operative microdialysis, it is usually hoped that there will be minimal tumor left to sample following surgery, often preventing sampling from multiple regions of the glioma. Intra-operative microdialysis can enable sampling of diverse regions within the live extracellular glioma microenvironment during standard-of-care glioma resections. However, low molecular weight catheters preclude collection of most extracellular analytes; slow flowrates combined with short intra-operative sampling times prevent acquisition of enough microdialysate volumes to enable multi-omic analyses. We hypothesized that we could safely and feasibly deploy intra-operative high molecular weight (HMW, 100kDA) microdialysis using an elevated perfusion rate of 2 µL/min^15^ to recover a usable volume of microdialysate containing diverse analytes, including metabolites, during a standard-of-care surgery. To that end, we initiated our intra-operative microdialysis trial in an initial cohort of five patients. To evaluate the feasibility of the intra-operative microdialysis approach, we aimed to measure microdialysate levels of the well-characterized and dialyzable oncometabolite, D-2-hydroxyglutarate (D-2-HG), known to be produced by IDH-mutant gliomas^23^. As such, patients were enrolled whose tumors could plausibly be IDH-mutant. Subsequent pathology revealed that three of these initial five patients indeed had IDH-mutant gliomas (Oligo^2^, Oligo^3^1, and Astro^4-mut^1); two patients were found to have IDH-wild type (WT) glioblastomas (GBM^WT^1, GBM^WT^2) (**Table 1**). To determine if D-2-HG levels in intra-operatively acquired microdialysate reflected IDH status, we utilized 10 µL of a 40 µL microdialysate sample from each catheter to quantify D-2-HG and L-2-HG via targeted metabolomic analysis. D-2-HG was elevated in microdialysate from IDH-mutant as compared to IDH-wild-Type (WT) tumors (median: 27.29 versus 0.63 µM) (**Supplementary Fig. S2A**), levels of L-2-HG were not different between groups (**Supplementary Fig. S2B**). Patient Astro^4-mut^1 yielded a >1000x difference in D-2-HG between non-enhancing tumor (FLAIR) and brain adjacent to tumor (831.80 versus 0.68 µM). These data demonstrated the feasibility of intraoperative HMW microdialysis to sample a dialyzable oncometabolite during standard-of-care surgery for tumor resection.

Since IDH status has been reported to impact the metabolism of IDH-mutant gliomas, we further analyzed these samples via untargeted metabolomic analyses--an increasingly leveraged strategy to gain more comprehensive insights regarding the global metabolome. We utilized 20 µL of microdialysate from each catheter for untargeted metabolomic analysis via the Metabolon platform^24^. Among hundreds of other metabolites, untargeted analysis reported total 2-HG. When plotted against the combined stereoisomer (D-2-HG + L-2-HG) levels from targeted analyses, a robust correlation (R^2^=0.9989) between the two platforms was observed across a 2,500-fold concentration range (**Supplementary Fig. S2C)**. Although limited sample volume precluded repeating this approach for other metabolites, these data supported the potential for untargeted metabolomic analyses to generate at least relatively quantifiable data.

Analysis of 2-HG peak area in a second cohort comprising 5 IDH-mutant and 5 IDH-WT cases, including one patient’s repeat surgery, confirmed increased levels in IDH-mutant tumors when compared to IDH-WT tumors (21.41x average normalized peak area in IDH-mutant tumor catheters compared to IDH-WT tumor catheters) (**Fig. 1C**). However, one patient with an unusual right frontal H3K27M-mutant infiltrative grade 4 astrocytoma also demonstrated moderately elevated 2-HG levels despite Next Generation Sequencing-confirmed IDH-WT status. This one apparent “outlier” illustrates that elevated extracellular 2-HG is not infallibly specific for IDH-mutant gliomas and is consistent with prior reports of occasionally elevated 2-HG in IDH-WT gliomas^25^.

Additionally, in two patients with grade 4 IDH-mutant astrocytomas, the extracellular abundance of 2-HG was several-fold less in enhancing than non-enhancing tumor (**Fig.1D**, FC (E/NE): 0.017x and 0.20x for patients Astro^4-mut^1 and Astro^4-mut^2, respectively). This observation raised the possibility that 2-HG could be lost down its concentration gradient into systemic circulation. Indeed, elevated 2-HG was previously reported in venous outflow of IDH-mutant high-grade gliomas^26^. An alternate explanation could be that enhancing tumor regions from IDH-mutant gliomas generate less 2-HG due to changes in the IDH metabolism of the tumor core. Arguing against this alternate explanation, a third patient with an enhancing grade 4 IDH-mutant astrocytoma showed enhancement throughout the lesion, precluding placement of a catheter in non-enhancing tumor. Although both tumor catheters were placed within the margins of contrast-enhancing tumor, the second tumor catheter targeted a region of less intense enhancement, suggestive of poorer central perfusion or relative necrosis (E†). Despite its placement in presumably poorly perfused tumor core, this catheter recovered 12.1-fold more 2-HG than the catheter in the most avidly contrast-enhancing region, suggesting that IDH-mutant metabolism was still present in the tumor core. Collectively, data from these three patients suggested that 2-HG may be lost from the extracellular compartment in regions of contrast-enhancing tumor due to increased diffusion across a disrupted blood-brain barrier (**Fig. 1D)**.

### Clustering of catheters: correlation analysis of the extracellular global metabolome

Having established that intra-operative HMW microdialysis could detect the relative abundance of an established oncometabolite of IDH-mutant gliomas, we next sought to evaluate the global extracellular metabolome across our patients’ 44 catheters. To ensure comparison and reproducibility of metabolite levels across patients, we analyzed 162 named metabolites that were present in at least 90% of the catheters (≥40/44 catheters). We first asked how patient’s catheters correlated to each other to identify possible patterns in the global metabolome based on patient identity, radiographic location, IDH mutation, and primary vs. recurrent status. Spearman correlation of all 44 catheters revealed that all samples from each patient with an oligodendroglioma (n=3 patients, 9/9 catheters) clustered based on patient identity. Conversely, 9/10 catheters from enhancing regions of WHO grade 4 astrocytomas clustered together (**Fig. 2A)**. Clustering of non-enhancing tumor regions was inconsistent, with some catheters clustering near the patient’s own enhancing catheters (patients Astro^4-mut^1, Astro^4-mut^2, and Astro^4-mut^3), while others clustered with brain catheters (**Fig. 2A)**, particularly those from a completely non-enhancing tumor (MolecGBM^WT^1). Although Patient GemAstro^3-mut^’s tumor did have enhancing components, these areas were extremely fibrous, hampering catheter placement. As such, both catheters were placed into non-enhancing regions. Interestingly, however, this patient’s catheters clustered with the enhancing tumor catheters, potentially suggesting flow of metabolites from nearby enhancing tumor regions (**Fig. 2A)**. Contrary to our expectations based on recently published tissue metabolomic studies, no obvious clustering was observed based on IDH status^27^, nor based on recurrence. Indeed, one patient’s enhancing catheter from their primary resection (GBM^WT^2: Catheter X) was most strongly correlated with the enhancing catheter from their repeat resection (GBM^WT^2B: Catheter X). In sum, results of correlation analyses suggest that the extracellular metabolome of molecularly diverse contrast-enhancing astrocytomas clustered by catheter location, whereas that of oligodendrogliomas clustered by patient. Perhaps unsurprisingly, the extracellular metabolome of non-enhancing tumor regions of WHO grade 4 astrocytomas appeared to exist on a spectrum, appearing in some cases more similar to the enhancing tumor and in other patients, closer to that of the “brain” catheter. However, visual inspection of relative metabolite abundance as a function of catheter demonstrated substantial heterogeneity across patients within each region (**Fig. 2B**).

### A reproducible metabolome of enhancing glioma versus brain

Given the substantial metabolic heterogeneity between patients, we asked if each patient could serve as their own control to identify the extracellular tumor metabolome. As the Spearman correlation had suggested clustering of enhancing tumor catheters (**Fig. 2A**), we began by comparing catheters in enhancing tumor as compared to adjacent brain. For each patient, metabolites were ranked from highest to lowest fold change based on metabolite peak area in enhancing tumor (E) versus brain adjacent to tumor (B). Although the magnitude of these fold changes varied between patients (**Supplementary Fig. S3)**, the rank of enhancing tumor versus brain-associated metabolites appeared similar in 3/3 patients with enhancing tumors from our initial cohort (**Supplementary Fig. S4A)**. We used the average ranks from these 3 patients to then evaluate the reproducibility of findings in the second cohort of 5 new patients and one patient’s recurrent (GBM^WT^2B) disease 10 months later. The ranked order of metabolites in the enhancing versus brain comparison appeared visually similar for all patients in both cohorts when ranked based on the average of the 9 cases **(Fig. 3Ai)**. Additionally, paired enhancing versus non-enhancing tumor catheters were available from 7 patients. Similar rank-based analyses were performed for enhancing versus non-enhancing tumor which revealed a surprisingly conserved rank of enhancing-to-non-enhancing metabolites across patients (**Fig. 3Bii; Supplementary Fig. S4B**). Conversely, the distribution of non-enhancing catheter-associated metabolites appeared less consistent (**Fig. 3Ci; Supplementary Fig. S4C**). Despite this diversity, when this non-enhancing-to-tumor metabolite distribution was queried in five patients from whom only non-enhancing tumor and brains catheters were available, a similar ranked metabolome was observed in two patients, Oligo^3^1 and GemAstro^3-mut^ (**Supplementary Fig. S5)**. These data demonstrated that metabolic diversity exists among patients with oligodendrogliomas and suggests that there could be recurrent metabolic patterns in some but not all non-enhancing tumors.

To more rigorously and quantitatively compare metabolic phenotypes between catheters and between patients, we utilized enrichment analysis (EA) to statistically identify similarities between and across patients’ catheters. Although more commonly applied to gene-based analyses, we repurposed this for metabolomic analysis. Using EA, we selected the top and bottom 35 metabolites based on fold change from each patient’s tumor vs. brain or enhancing vs. non-enhancing tumor ranked metabolite list (“metabolite sets”) (**Supplementary Fig. S1**). We then determined where these metabolites fell in the ranked metabolite list of tumor versus brain or enhancing vs. non-enhancing tumor for the remaining patients. “Positive enrichment” meant that the metabolites fell at the tumor-end of the ranked list, while “negative enrichment” indicated that they fell at the brain-end of the ranked list. In patients with enhancing astrocytomas, each patient’s individual enhancing tumor signature could be robustly and positively identified in the enhancing tumor of the other patients, suggesting a convergent glioma metabolome (**Fig. 3Aii)**. Conversely, each patient’s brain signature could be found at the “brain” end of every other patients’ ranked metabolite list **(Fig. 3Aii)**. Leveraging these individual patient-level data, these analyses proved highly robust across patients with a False Discovery Rate (FDR) of 0.000 for most interpatient comparisons of enhancing tumor versus brain metabolites (**Fig. 3Aii, Supplementary Table 2)**. For independent reference, the previously published post-operative tumor versus adjacent brain microdialysate metabolome from Björkblom et al^17^ was evaluated and found to be present in the enhancing catheters of astrocytomas (FDR of 0.002 or less, Supplementary Table 2). Indeed, among 24 reported brain- or tumor-associated metabolites, all but one metabolite revealed a reproducible pattern in our data set (**Supplementary Fig. S6**). These data confirmed the robust nature of the extracellular enhancing glioma metabolome, being readily and comparably discernible in both the intra-operative and post-operative settings.

Enrichment analyses of enhancing versus non-enhancing catheters again demonstrated that enhancing catheters were metabolically separate from their non-enhancing catheter counterparts and that this could be consistently found across most patients (most FDRs=0, except GBM^WT^1, **Fig. 3Bii;** Supplementary Table 3). As expected from Figure 3Ci, non-enhancing catheters were rarely enriched for each other when compared to brain (**Fig. 3Cii**, Supplementary Table 4). Overall, these data suggest that the extracellular metabolome of enhancing glioma is distinct from, and more consistent than that of non-enhancing glioma.

### Metabolites of the enhancing and non-enhancing glioma extracellular microenvironments

After demonstrating that enhancing astrocytomas exhibit a conserved extracellular metabolome via rank-based and formal enrichment analyses (**Fig. 4A)**, we next asked which metabolites define this extracellular microenvironment across the full cohort of all 9 surgeries. The Wilcoxon rank sum test for non-parametric distributions, paired enhancing vs. brain catheters revealed 48 significantly elevated metabolites in the enhancing tumor catheter and 22 significantly elevated metabolites in the brain catheter using cutoff criteria of p-value≤0.05 and FC≥2 (**Fig. 4A; Table 2)**. The top enhancing astrocytoma metabolite was guanidinoacetate (GAA) (E/B = 126.32x), which, to our knowledge, has not previously been reported in the context of glioma. The enhancing glioma signature was also defined by greater relative abundance of proline (FC=10.18x), carnitine-family metabolites (deoxycarnitine, carnitine, and acetylcarnitine at FC= 8.71x, 7.65x, and 7.47x, respectively), and N1-methyl-2-pyridone-5-carboxamide (2-PY) (FC=7.18x) (all at p<0.005). Metabolites defining the brain metabolome included arabitol (FC= 0.04) and arabonate (FC=0.08), in addition to canonical brain-associated metabolites, such as N-acetylaspartate (NAA)^28^ (all p<0.005).

We then asked if these metabolites were unique to enhancing tumor when compared to non-enhancing tumor for each patient (**Fig. 4B)**. Again using a Wilcoxon rank sum test for non-parametric distributions, twenty-eight metabolites were significantly higher in the enhancing glioma metabolome, while twenty metabolites were greater in the non-enhancing glioma metabolome (**Table 2**). Interestingly, 27/28 enhancing glioma metabolites, had also been found in the enhancing glioma-vs-brain comparison, again demonstrating a robust extracellular metabolome of enhancing glioma (relative abundance of representative metabolites shown in **Fig 4C**). Of the twenty non-enhancing versus enhancing glioma metabolites, 15 overlapped with brain (versus enhancing tumor). These data suggest the continued presence of brain-associated metabolites within non-enhancing portions of glioma. Indeed, when non-enhancing glioma catheters were compared to brain **(Fig. 4D)**, no metabolites were unique to the brain metabolome, while fifteen metabolites were higher in non-enhancing tumor, including GAA (FC NE/B=13.63x). Overall, these data demonstrate that a specific set of metabolites define the extracellular microenvironment of enhancing HGGs when compared to non-enhancing glioma and brain adjacent to tumor.

### Plasma in the glioma extracellular metabolome

By definition, contrast-enhancing glioma regions have a more disrupted blood-brain barrier than non-enhancing regions^29^, facilitating solute diffusion between the intravascular compartment and tumor microenvironment. Our data demonstrated a similar metabolic signature of enhancing, but not non-enhancing portions, of gliomas when compared to brain (**Fig. 3A, C)**, and metabolic differences between enhancing vs. non-enhancing portions of gliomas (**Fig. 3B, 4B**). As such, we hypothesized that enhancing glioma metabolites may originate from plasma preferentially entering regions of greater BBB disruption where there is decreased resistance to diffusion. To test this hypothesis, we utilized a ranked metabolite list of “bloodiness” from identically paired clean versus bloody CSF samples (see methods). In the 7 patients for whom paired enhancing and non-enhancing microdialysates were obtained, we first compared the relative enrichment of enhancing versus non-enhancing brain catheters for bloody versus clean CSF using each patient’s ranked enhancing versus non-enhancing list (Cath. X/Y). Enrichment analyses demonstrated significant enrichment in 7/7 patients for at least one of (i) enhancing tumor enriched in bloody CSF, or (ii) non-enhancing tumor enriched in clean CSF; 5/7 patients showed significant enrichment in both analyses (FDR<0.05) (**Fig. 5A**). Evaluation of specific enhancing versus non-enhancing tumor-associated metabolites from Fig 4B revealed higher abundance of bloody CSF-associated metabolites in enhancing tumor (**Fig. 5B**). As expected, performing enrichment analyses on enhancing glioma versus brain ranked list again demonstrated that enhancing glioma catheters were enriched for bloody CSF in 9/9 patients **(Supplementary Fig. S7)**. Results in non-enhancing tumor suggested a modest enrichment of bloody CSF-associated metabolites in some catheters, perhaps suggestive of either some sub-radiographic BBB disruption^29^, or metabolite diffusion from adjacent enhancing regions. Collectively, these data suggest that plasma-derived metabolites from a disrupted BBB significantly contribute to the extracellular metabolome of enhancing gliomas **(Fig. 5C)**.

**Figure 5.**
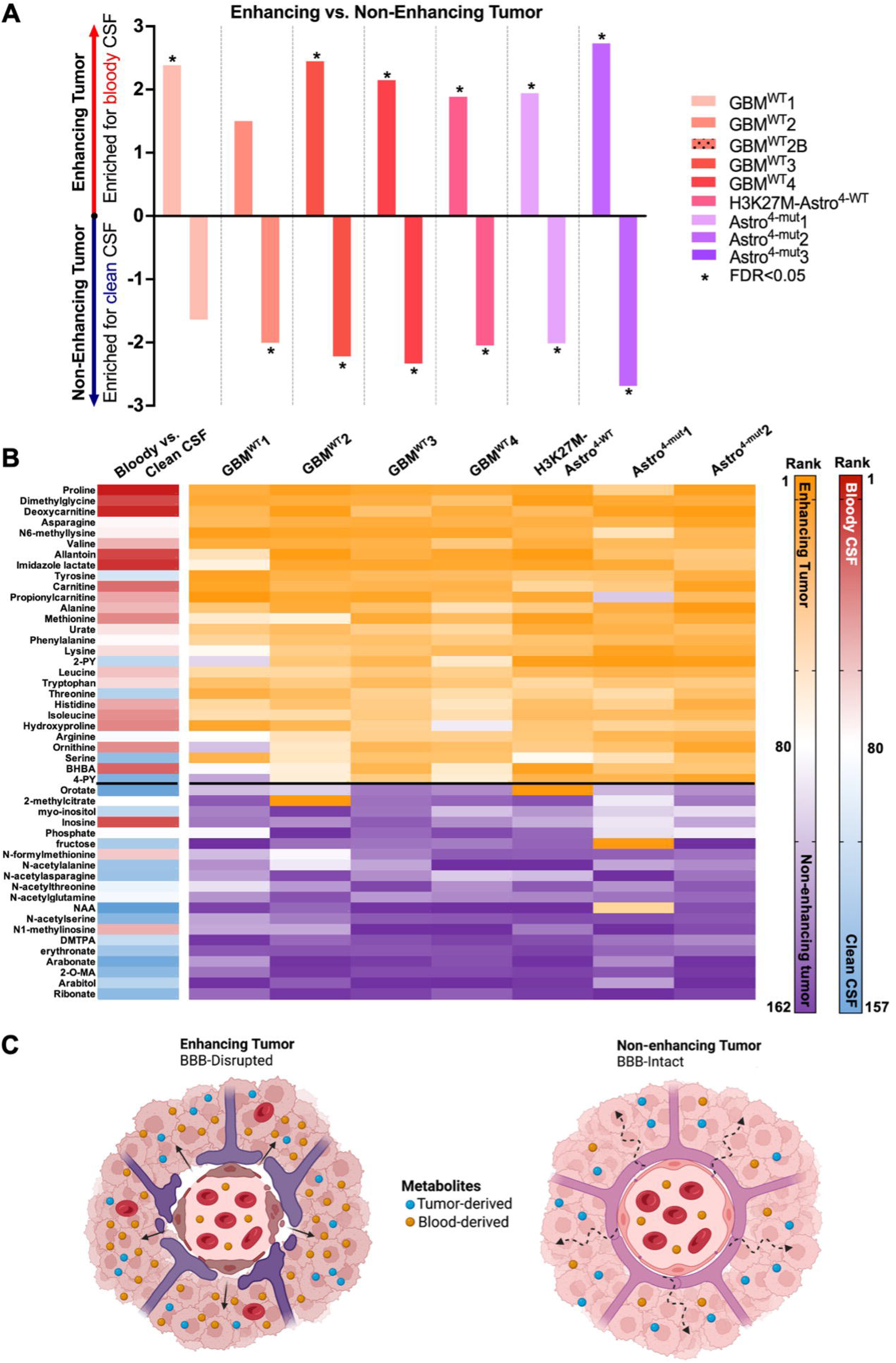
Enhancing glioma microdialysate is enriched for plasma-derived metabolites. **(A)** Enrichment analysis was utilized to determine the enrichment of each patient’s enhancing vs. non-enhancing tumor catheters (ranked list: Cath. X vs. Y) for high-plasma versus low-plasma CSF (metabolite sets). Positive normalized enrichment scores (NES) indicate metabolic similarities to high-plasma CSF; negative NES indicate metabolic similarities to low-plasma CSF (* = FDR≤0.05). (**B)** The rank of each metabolite from the significant enhancing versus non-enhancing glioma metabolite in Figure 4B is shown for each patient (E: purple, rank 1, to NE: orange, rank 162), along with the metabolite’s rank in a bloody vs. clean CSF ranked fold-change list (bloody: red, 1 – clean: blue, 157). (**C)** Proposed model depicting the impact of blood-brain barrier disruption on the enhancing versus non-enhancing glioma extracellular metabolomes (created using BioRender).

One recent intraoperative study compared metabolites within the glioma’s arterial versus venous supply to determine which were consumed versus produced by the gliomas^26^. We analyzed their raw data maintaining thresholds we had used for our own data (FC≥2; p≤0.05), and initially found no significantly differentially abundant metabolites. By removing the FC criterion, 4 significant metabolites were identified. Taurine was more abundant in venous than arterial glioma blood, suggesting production by the tumor. In agreement with this result, we found taurine to be significantly more abundant in both enhancing and non-enhancing tumor as compared to brain (FC E/B= 2.99x, NE/B = 2.08x). Three metabolites were more abundant in glioma arterial than venous blood, suggesting utilization by the tumor **(Sup. Fig. S8)**. In agreement with our data, one of these, alanine, was part of our enhancing glioma metabolome (**Table 2**) and enriched for bloody CSF **(Fig. 5B)**, consistent with the hypothesis of BBB disruption improving the glioma’s access to needed metabolites. One limitation of the arterial-venous sampling study was that no parallel analyses were performed to compare metabolite utilization and production in adjacent brain. For example, although glucose was identified as utilized by “glioma” in their data, glucose readily crosses easily crosses the BBB and is highly utilized by brain, making fluorodeoxyglucose (FDG)-positron emission tomography (PET) unhelpful to discriminate tumor from brain^30^. Indeed, extracellular glucose was not differentially abundant in our tumor versus brain microdialysate. Nevertheless, these limited available data further support our hypothesis that the enhancing glioma metabolome is significantly enriched for metabolites from peripheral blood, at least some of which directly support glioma metabolism. Conversely, extracellular metabolites that are differentially abundant in both enhancing and non-enhancing tumor when compared to brain are most likely generated by tumor cells, including GAA and taurine.

## DISCUSSION

We utilized an intraoperative window-of opportunity to perform microdialysis, sampling the extracellular metabolome of radiographically distinct regions of human gliomas *in situ*. Findings revealed that 1) 2-HG is robustly elevated in IDH-mutant gliomas, with potentially highest levels in non-enhancing tumor regions. 2) GAA is a previously unreported extracellular glioma metabolite that highly differentially abundant in enhancing (126.32x) and non-enhancing (13.63x) glioma as compared to brain; 3) The extracellular metabolome of enhancing glioma is conserved across patients, comprises largely amino acids and carnitines, and is enriched for plasma-derived metabolites.

Acute intraoperative microdialysis yielded relative levels of hundreds of metabolites for untargeted metabolomics analyses, including 2-HG. IDH-mutant gliomas epitomize the oncogenic relevance of altered metabolism^31,32^. We observed elevated 2-HG in the microdialysate of IDH-mutant tumors via both targeted (D vs L-2-HG) and untargeted (total D+L-2HG) analysis (**Fig. 1C; Supplementary Fig. 2)**. Total 2-HG was previously used as a pharmacodynamic biomarker for IDH inhibitors in tissue^33^ and via magnetic resonance spectroscopy^34-36^. As microdialysis can be utilized for pharmacokinetic studies of dialyzable drugs^9^, the ability to quantify extracellular 2-HG *in vivo* across a >1000x concentration range **(Fig. 1C)** suggests that extracellular 2-HG within microdialysate could provide an pharmacodynamic readout for relevant early phase clinical trials^37^. Interestingly, 2-HG was 5-12x lower in the most avidly enhancing tumor region (n=3) (**Fig. 1D)**, suggesting that some tumor metabolites may be lost down a disrupted blood brain barrier—a consideration of potential importance for longitudinal pharmacodynamic monitoring of tumor interstitial fluid.

Importantly, although IDH-mutations are thought to drive early tumorigenic metabolic and epigenetic changes^32,38^, the global extracellular metabolomes of enhancing IDH-mutant versus IDH-WT grade 4 astrocytomas were otherwise indistinguishable (**Fig. 2A, 3A)**. Recently published work from over 200 glioma tissues demonstrated less separation between IDH-mutant grade 4 astrocytomas from GBMs^27^, than lower grade IDH-mutant gliomas, potentially suggesting metabolic convergence of high-grade gliomas. Greatest similarities were seen across patients in the extracellular metabolome of contrast-enhancing tumors, which could be expected if improved availability of plasma-associated metabolites impacts tumor metabolism.

Interestingly, in each of four patients with non-enhancing oligodendrogliomas the 3 catheters clustered together based on patient identity (**Fig. 2A)**. Such close and consistent clustering of catheters within each of these patients could reflect the frequently more diffuse nature of oligodendrogliomas, with each catheter sampling an admixture of metabolically individualized tumor and brain.

Cancer-associated metabolic vulnerabilities are of increasing interest as therapeutic targets. In contrast to gliomas’ notorious genomic heterogeneity^39,40^, a more focused inventory of metabolic strategies may be utilized to combat bioenergetic demands^41^ oxidative stress^42^, hypoxia^43^, and nutrient deprivation^44,45^. Our rank-based analyses revealed an enhancing glioma extracellular metabolome that was consistent across primary and recurrent IDH-WT and mutant lesions, as well as an H3K27M-mutant tumor (**Fig. 3A(i-ii)**). This signature appeared comparable to that previously obtained in the context of post-operative microdialysis^16,17^, consistent with reproducibility of the enhancing glioma metabolome across clinical and technical variables (**Supplementary Table 1**), research teams and contexts spanning intra-operative versus post-operative sampling. However, our data demonstrate for the first time that most of the metabolites in the enhancing glioma extracellular metabolome are absent from the non-enhancing glioma metabolome. These results suggest that the extracellular metabolome of enhancing gliomas may be largely defined by metabolites from systemic circulation that more easily access the tumor microenvironment through a disrupted BBB. Although catheter insertion may hypothetically result in some degree of BBB disruption, intra-patient catheter comparisons revealed significant enrichment for plasma-associated metabolites, suggesting that the extent BBB disruption induced from catheter insertion was negligible compared to the tumor-induced disruption in enhancing versus non-enhancing tumor. Furthermore, our findings are in alignment with the observation that aggressive portions of gliomas avidly uptake amino acids from the systemic circulation, serving as the basis for highly sensitive and specific glioma PET tracers, including tyrosine^46^ and methionine,^47^ both of which were more abundant in enhancing glioma than non-enhancing tumor or brain **(Fig. 4; Table 2)**.

Given the established importance of multiple identified plasma- and enhancing tumor-associated metabolites, including amino acids and carnitines to cancer biology^48-50^, BBB disruption itself may underlie the metabolic similarities across molecularly diverse HGGs by providing a continuous source of protumorigenic metabolites. Anecdotally, our patient with a completely non-enhancing molecular GBM had evidence of this lesion on a CT obtained 5 years prior to diagnosis—consistent with a lower rate of growth than patients with enhancing lesions. Indeed, the adverse prognostic impact of a contrast-enhancement in gliomas, including histologically low-grade tumors, is well recognized^51,52^, as is the positive impact on survival of gross total resection of enhancing tumor across all molecularly defined GBM subgroups and ages^53^. As such, our findings suggest that blood brain barrier disruption itself may transform the extracellular glioma metabolome into a wellspring of nutrients for accelerated growth, particularly in enhancing tumor regions.

At this point, we cannot definitively answer the “chicken” or “egg” question of whether BBB disruption occurs for the purpose of accessing systemically-derived analytes or if it is simply a pre-existing process that a glioma can utilize to its advantage. Many of the plasma-associated metabolites enriched in enhancing tumor have been reproducibly associated with cancer biology^54^, including the amino acids proline^48^, glycine^55^, and amino acid derivatives, such as N6-methyllysine. Similarly, carnitines (**Fig. 4A-B, D(iii-iv)**) have been linked to energy production via fatty acid oxidation in glioma^56,57^. It is possible that the induction of blood-brain barrier disruption by proliferating gliomas^58^ may represent an adaptive response to nutrient deprivation observed in gliomas^59-61^, catalyzing a feedforward cycle of nutrient utilization and recruitment. Mechanistic studies selectively manipulating BBB permeability or extracellular plasma abundance will be required to dissect these interactions. If our hypothesis of disrupted BBB-mediated tumor aggressiveness is correct, maintaining and restoring BBB integrity may prove an independently important therapeutic goal, distinct from angiogenesis-blocking agents such as bevacizumab^62^.

Although we used UPLC-MS/MS to maximize our yield of potentially novel glioma metabolites, we had not expected to find a new extracellular glioma metabolite that could outperform all other enhancing glioma metabolites by over 10-fold. Although GAA was present at a low abundance in bloody CSF and not detected in clean CSF, its disproportionately high fold change in both enhancing and non-enhancing tumor versus brain (126.32x, and 13.63x, respectively; Figure 4A, D(i), **Table 2**) suggest it may be produced by the tumor. No CNS efflux transporter exists for GAA at the BBB^63^. As such, if generated as a result of glioma metabolism, it is liable to accumulate. GAA also accumulates in patients with GAMT deficiency, leading to impaired creatine production, seizures, and cognitive impairment^64^. GAA is a precursor for creatine, levels of which were slightly elevated in non-enhancing (2.25x; p=0.03), but not enhancing tumor (**Table 2**). Interestingly, GAA is co-produced with ornithine, which was slightly higher in enhancing tumor when compared to non-enhancing tumor (2.36x, p=0.03) and brain (3.07x, p=0.004) (**Table 2**). Ornithine is the substrate for polyamine synthesis via ornithine decarboxylase (ODC) which is upregulated in cancers, including glioblastoma, to support catabolic demand, regulate intracellular pH in the acidic tumor core, and protect against immune surveillance^65^. It is therefore tempting to speculate that increased GAA could be a byproduct of increased glioma polyamine synthesis and utilization. If true and based on the substantial fold-change, extracellular GAA could serve as a biomarker of not only local glioma abundance but also pharmacodynamic response to ODC blockade. ODC blockade with Difluoromethylornithine (DFMO) remains in clinical trials for glioma (NCT02796261); though upregulation of polyamine transporters may mediate metabolic compensation leading to improved survival with dual blockade of ODC and polyamine transporters (via AMXT-1501), in preclinical models of diffuse midline gliomas and other cancers^66,67^. To determine if GAA can be used as a dynamic biomarker of ODC production *in situ*, we are opening a post-operative microdialysis study under an IND, monitoring microdialysate levels of GAA in unresectable regions of residual tumor with or without DFMO +/- AMXT1501. Further dynamic insights may be achievable with isotope tracing and are under development.

## Conclusions

With the help of our patients, and intraoperative microdialysis we have shown that GAA is surprisingly abundant in the glioma microenvironment, and that plasma-associated metabolites largely define the enhancing glioma microenvironment, potentially positioning them to support glioma aggressiveness. Our patients understand the formidable odds of their disease. We present their data as a first installment toward what we intend to be a growing collaborative, open-source repository of individualized data from live human gliomas. We (both patients and investigators) hope that the data and insights shared can help inspire novel translational paradigms. By leveraging neurosurgical access to live human disease, we hope to accelerate tangible progress within the lifetimes of individual patients.

## Data Availability

The data we have collected will be made publicly available upon later submission.

## Acknowledgements

We would like to thank each of our patients who selflessly participated in this study and without whom this work and any advancement in the field of neuro-oncology would not be possible. We also thank Elizabeth Oishi for regulatory support and the Mayo Clinic Neurosurgery Clinical Research team, and the Mayo neurosurgery operative staff for their invaluable support.

## List of Abbreviations

2-HG: 2-hydroxyglutarate
2-PY: N1-methyl-2-pyridone-5-carboxamide
B: brain
BBB: Blood-brain barrier
CNS: central nervous system
CSF: cerebrospinal fluid
CT: computed tomography
DFMO: difluoromethylornithine
E: enhancing tumor
FC: fold-change
FDR: false-discovery rate
GAA: guanidinoacetate
GBM: glioblastoma
GSEA: gene set enrichment analysis
HGG: high-grade glioma
HMW: high molecular weight
IDH: isocitrate dehydrogenase
IND: investigational new drug
LC/MS: liquid chromatography-mass spectrometry
NAA: N-acetylaspartate
NE: non-enhancing tumor
ODC: ornithine decarboxylase
WT: wild type
UPLC-MS/MS: Ultra-performance liquid chromatography tandem mass spectrometry

