## Supplemental methods, figures, and tables for "Does Blood-Brain Barrier Disruption Define the Glioma Extracellular Metabolome?"

**Supplementary Material (Supplemental Methods, Figures, Legends and Tables)**

**Supplementary Methods**

**Intraoperative Microdialysis**

Each patient underwent intraoperative microdialysis using two to three M-dialysis 100 kDA catheters and variable rate microdialysis pumps (M-dialysis 107 Pump) under an investigational device exemption. Catheters were continuously perfused at 2 μL/min with isotonic perfusate containing 3% Dextran or albumin to balance oncotic pressure and maintain net fluid flux across the catheter membrane. Catheter target locations were selected based on pre-operative magnetic resonance imaging (MRI) by the surgeon to facilitate sampling of radiographically diverse regions, including contrast-enhancing, non-enhancing (T2 FLAIR hyperintense) tumor, and brain adjacent to tumor when possible and applicable. Unless otherwise specified, non-enhancing tumor areas were chosen that were outside not part of the necrotic tumor core. After reviewing the imaging at the time of surgery, catheter trajectories were planned for each catheter. The intended target depth was measured via the neuro-navigation system and then marked on the catheter with sterile adhesive tape (Steri-Strip).

After dural opening, but prior to tumor resection, a small pial opening was made at the intended catheter entry sites. Each pre-primed catheter was then advanced manually along pre-planned trajectories to the pre-determined depth with the aid of computer-guided neuro-navigation. Cortical surface mapping procedures were performed as needed after catheter implantation. Surgical resection then began, working initially away from the implanted catheters. Collection vials were initially changed after all catheters were in place and every 20 minutes thereafter to obtain multiple 40 μL aliquots per catheter. Collection continued until at least two aliquots had been collected. Each aliquot was immediately labeled and placed directly on dry ice in the operating room. The second aliquot after catheter placement was submitted for metabolomic analysis to minimize potential confounders of variably equilibrated microdialysate in the first aliquot. Any additional aliquots, when available, were saved for future analysis. Catheters were removed prior to resection of the sampled region of brain or tumor. With exception of intraoperative microdialysis, no other aspects of the surgical procedure, or post-operative care were altered. Extent of resection was guided in some cases by awake language and/or motor mapping and in all cases by imaging via computer-guided neuro-navigation. 5-aminolevulic acid (ALA) fluorescence was additionally used to guide resection of enhancing lesions and intraoperative ultrasound was used to guide extent of non-enhancing lesions. Microdialysate aliquots were stored at -80C until analyzed. Patient samples were analyzed in 2 cohorts **(Supp. Figure S4)**. Correlative clinical information was obtained from the medical record. No complications occurred that were attributable to use of intraoperative microdialysis. Patient GBM^WT^1 underwent a second surgery six months after the index operation for resection of a small focus of enlarging enhancement; microdialysis was not performed; pathology demonstrated only pseudo-progression; microdialysis . Patient GBM^WT^2 underwent a repeat resection for a recurrent enhancing lesion nine months after the primary resection; pathology revealed recurrent IDH-WT GBM (GBM^WT^2B). No attempt was made to correlate results to patient survival, since most patients remained alive at the time of manuscript submission.

**Pathology and Molecular Tumor analysis**

Tumor samples from each patient underwent standard histopathological analysis for diagnostic purposes as part of the routine clinical workflow. Immunohistochemical analysis for IDH R132H was used to determine IDH status. Negative cases were confirmed by IDH1/2 sequencing. GBMs were evaluated for O^6^-methylguanine-DNA methyltransferase (MGMT) methylation. Telomerase reverse transcriptase (TERT) promotor mutations were identified by sequencing. 1p/19q co-deletion, cyclin dependent kinase inhibitor 2A (CDKN2A) status, and karyotype complexity were evaluated via chromosomal microarray.

**Targeted Analysis of D/L-2-HG**

Targeted metabolomic analysis of microdialysate and CSF samples was performed by the Mayo Clinic metabolomic core facility. L and D isomers of 2-hydroxyglutaratic acid were separated and quantified by liquid chromatography mass spectrometry (LC/MS). Internal standard solution containing U-13C labeled 2-hydroxyglutarate (2-HG) was added to microdialysate. Proteins were removed by adding 500 μl of chilled 80% methanol solution to the sample mixture. The mixture was allowed to incubate on ice for 40 minutes prior to removal of the supernatant. After drying the supernatant in the speed vac under medium heat, the samples were derivatized with Diacetyl-L-Tartaric Anhydride (25 mg/ml in 4:1 dichloromethane: acetic acid). Samples were dried and resuspended in 100μL of water before analysis on a Nexera X2 UPLC module coupled with an AB Sciex Triple Quad 6500 mass spectrometer. Metabolites were separated on an Acquity HSS-T3, 1.8 μm, 2.1 x 50 mm column (Waters Corp, MA, USA), held at 40 °C, using 99% water, 1% acetonitrile, and 5 mM ammonium formate in water, adjusted to pH 3.3 with formic acid, as mobile phase A and 99%acetonitrile, 1% water, and 0.1% formic acid as mobile phase B. The flow rate was 0.3 mL/min and 2-HG D- and L- isomers were separated with an isocratic elution (99%A, 1%B) for 8 minutes. The mass spectrometer was operated in ESI- mode, monitoring mass transitions of m/z 363 -› 147 for DATAN labeled 2-HG and 147 -› 129 for 2-HG. Concentrations of both isomers were measured against a 10-point calibration curve that underwent the same derivatization. Using these methods, the limit of quantification was 20 nM with an inter-sample coefficient of variance of 11.30%. Human bone marrow plasma, as a biological reference, contains 0.52μM D-2-HG.

**Untargeted Metabolomic Analysis**

Untargeted metabolomic analysis was performed by Metabolon, Inc. Two fractions were evaluated with reverse phase (RP) UPLC-MS/MS with positive ion mode ESI, one by RP/UPLC-MS/MS with negative ion mode ESI and one with Hydrophobic interaction chromatography (HILIC) UPLC-MS/MS with negative ion mode ESI. Throughout the sample processing, multiple controls were analyzed with the experimental samples, including a pooled matrix sample, extracted water samples, and a cocktail of QC standards for instrument performance monitoring and chromatographic alignment. Relative standard deviation was measured for instrument variability control and overall process variability was determined by median relative standard deviation (RSD) for all endogenous metabolites. Raw data was extracted from UPLC-MS/MS results, peak-identified, and QC processed according to Metabolon standards. Metabolites were identified by compared data to Metabolon’s library which includes information on retention index (RI), mass to charge ratio (m/z), and chromatographic data of molecules.

Fifty-one microdialysate samples were analyzed, including 7 blanks comprising perfusate that had passed through catheters during the flush cycle prior to collecting the first aliquot. Forty-four microdialysate samples comprised the second aliquot collected from each catheter (3 catheters in 14 cases, 2 catheters in one case) after intraoperative placement within the tissue. Following protein removal, four fractions of the metabolite extract were randomly run across the platform and analyzed by ultra-performance liquid chromatography tandem mass spectrometry (UPLC-MS/MS). Further details can be found in Supplemental Methods. Ten samples were found by Metabolon to contain <20 μL. As such, identified peak areas for metabolites in this sample were scaled to 20 μL prior to use in analysis (indicated in Supplementary data). Batch-normalized peak areas were utilized, normalizing each metabolite to have a median of 1 in each of the three batches (identified in Supplementary Data).

**Supplementary Figures and Legends**

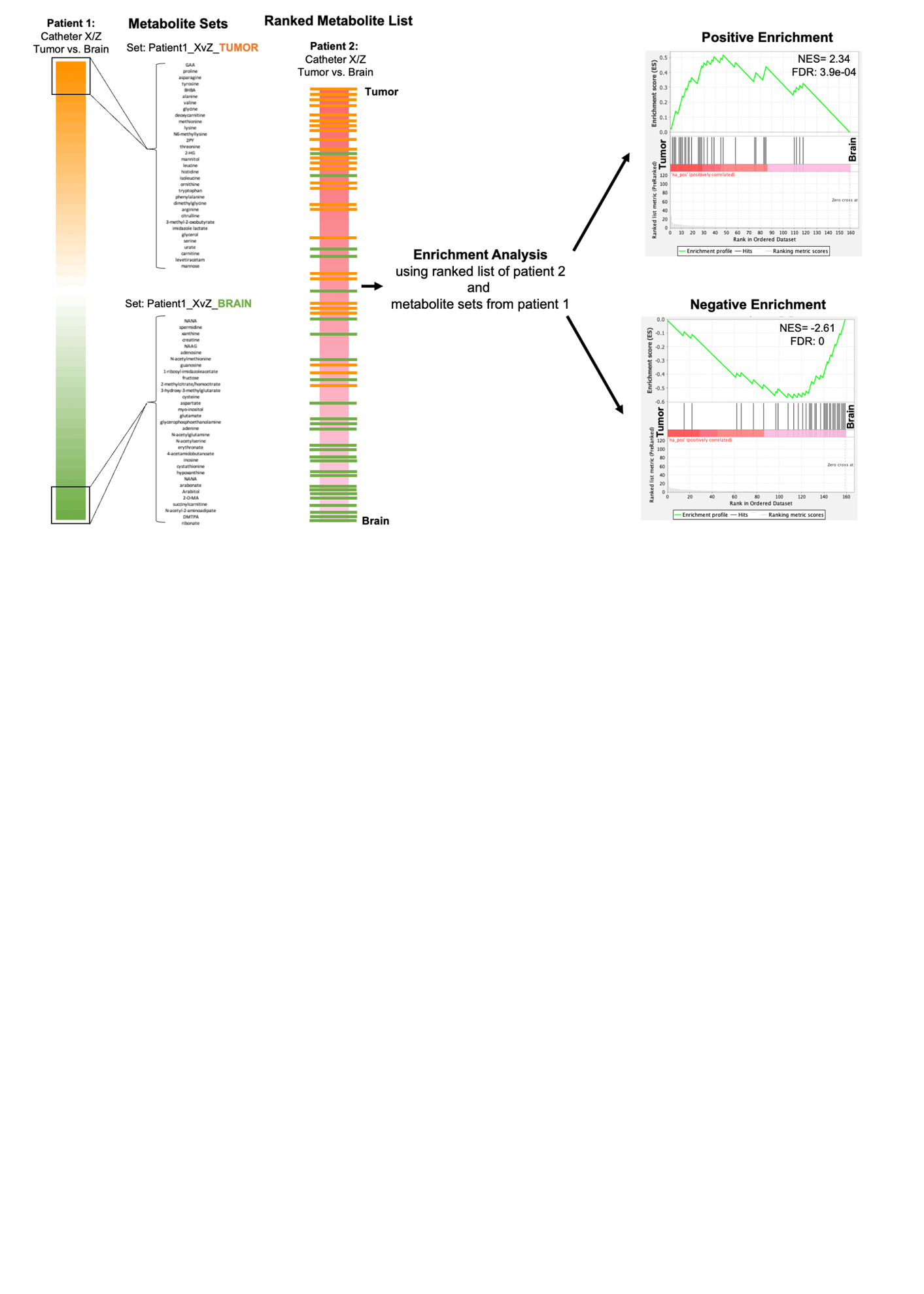

**Supplementary Figure S1. Graphical depiction of the repurposing Gene Set Enrichment Analysis (GSEA) for metabolomic rank-based analyses**. See methods for further details on how ranked lists and metabolite sets are generated.

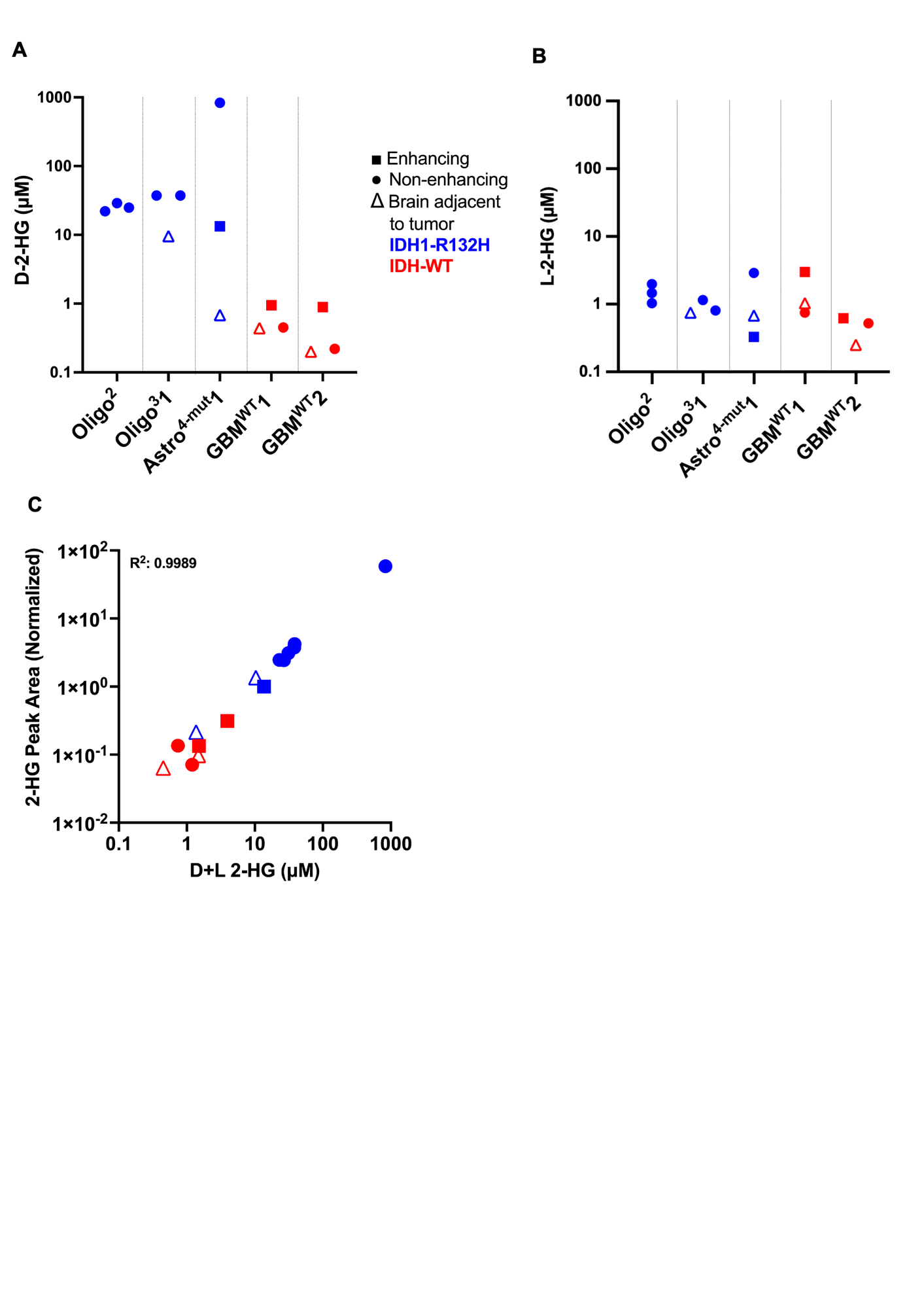

**Supplementary Figure S2**. **Targeted and untargeted analyses of 2-hydroxyglutarate in the discovery cohort. A)** D-2-hydroxyglutarate (D-2-HG) concentration, and (B) L-2-hydroxyglutarate (L-2-HG) as quantified via targeted metabolomics (LC-MS), is shown for microdialysate from each of the 15 catheters in the discovery cohort. Patient IDH status is indicated as mutant (blue) or wild type (red). Symbol shape indicates catheter placement location (see legend). (**C**) Untargeted metabolomic analysis was performed on 20 𝜇L of microdialysate from the same aliquot as used for targeted metabolomics in the discovery cohort. 2-HG peak areas obtained via untargeted metabolomic analysis do not discriminate between D-2- and L2-HG. These were correlated to the sum total of D-and-L-2-HG measured via targeted LC-MS.

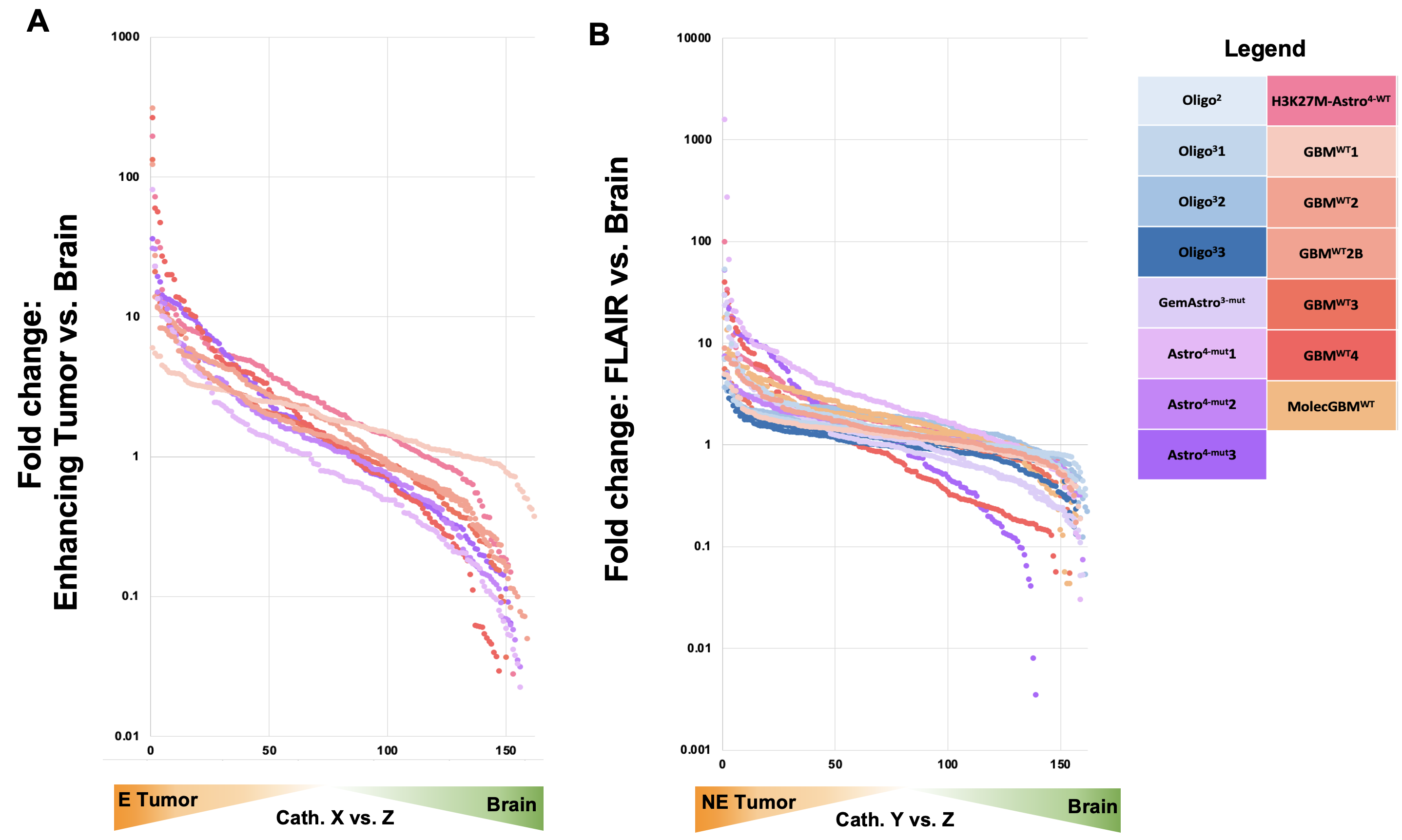

**Supplementary Figure S3**. **Fold-change values for each metabolite based on rank in Enhancing or Non-enhancing tumor versus brain.**

Scatter plots demonstrating the relationship between fold change and rank in catheters X versus Z (**A)** and catheter Y versus Z **(B)** across the 162 metabolites presents in at least 40/44 catheters for all patients.

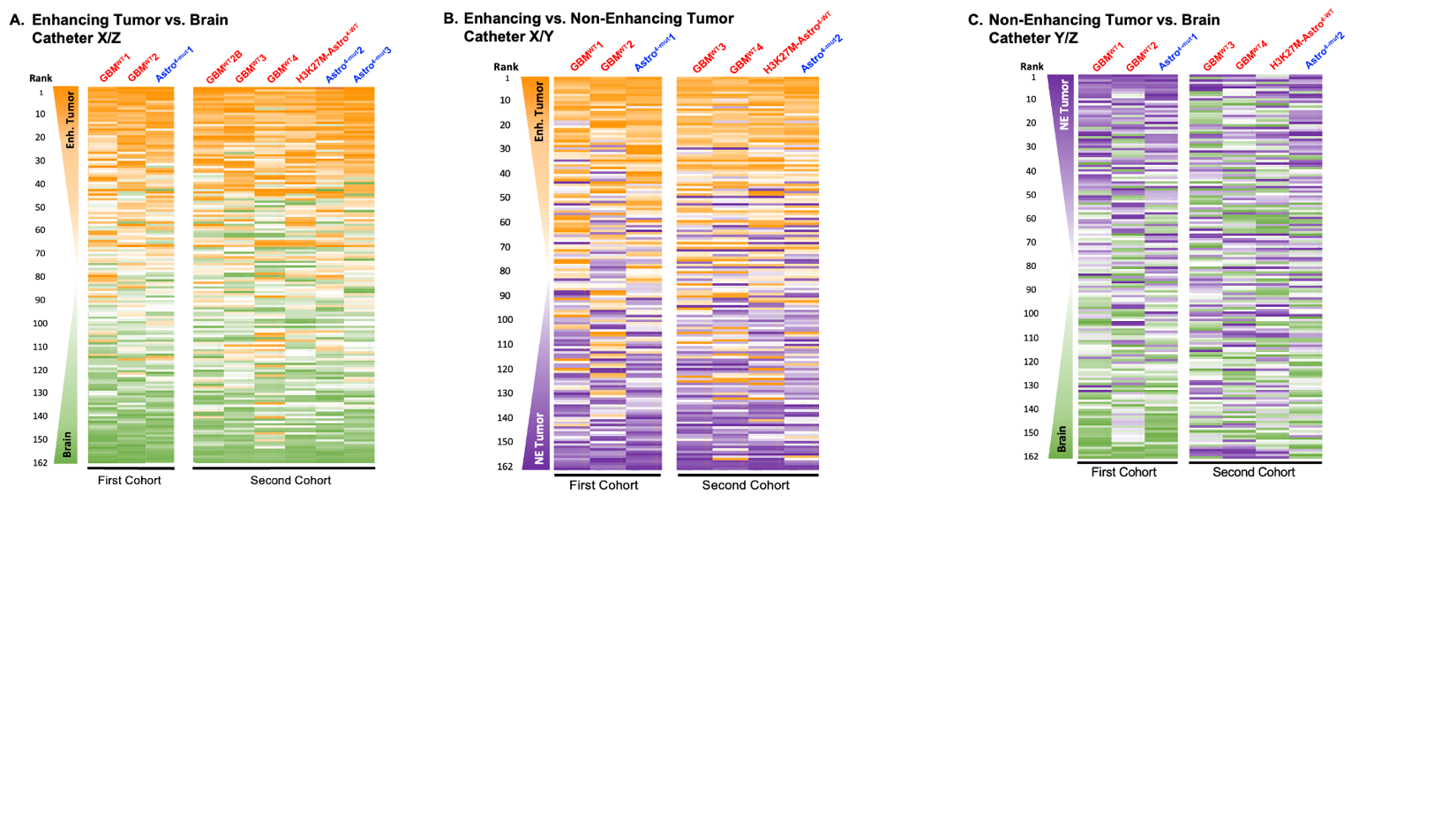

**Supplementary Figure S4. Metabolic signatures of the enhancing versus non-enhancing glioma versus brain metabolites, using first and second cohort-based analyses.**

162 metabolites present in at least 40/44 catheters were ranked according to the (A) enhancing tumor-versus-brain (X/Z), (B) enhancing versus non-enhancing tumor (X/Y), or (C) non-enhancing tumor versus brain (Y/Z) fold change in each patient. The rank order of each metabolite in each 2-catheter tumor/brain comparison (e.g., catheter X versus Z) is conveyed as a heat map from 1 to 162 (orange: enhancing tumor, purple: non-enhancing tumor, brain: green). Metabolites are listed based on the average of ranks of (A) enhancing vs. brain (B) enhancing vs. non-enhancing tumor, and (C) non-enhancing tumor vs. brain in the first cohort (Astro^4-mut^1, GBM^WT^1 and GBM^WT^2).

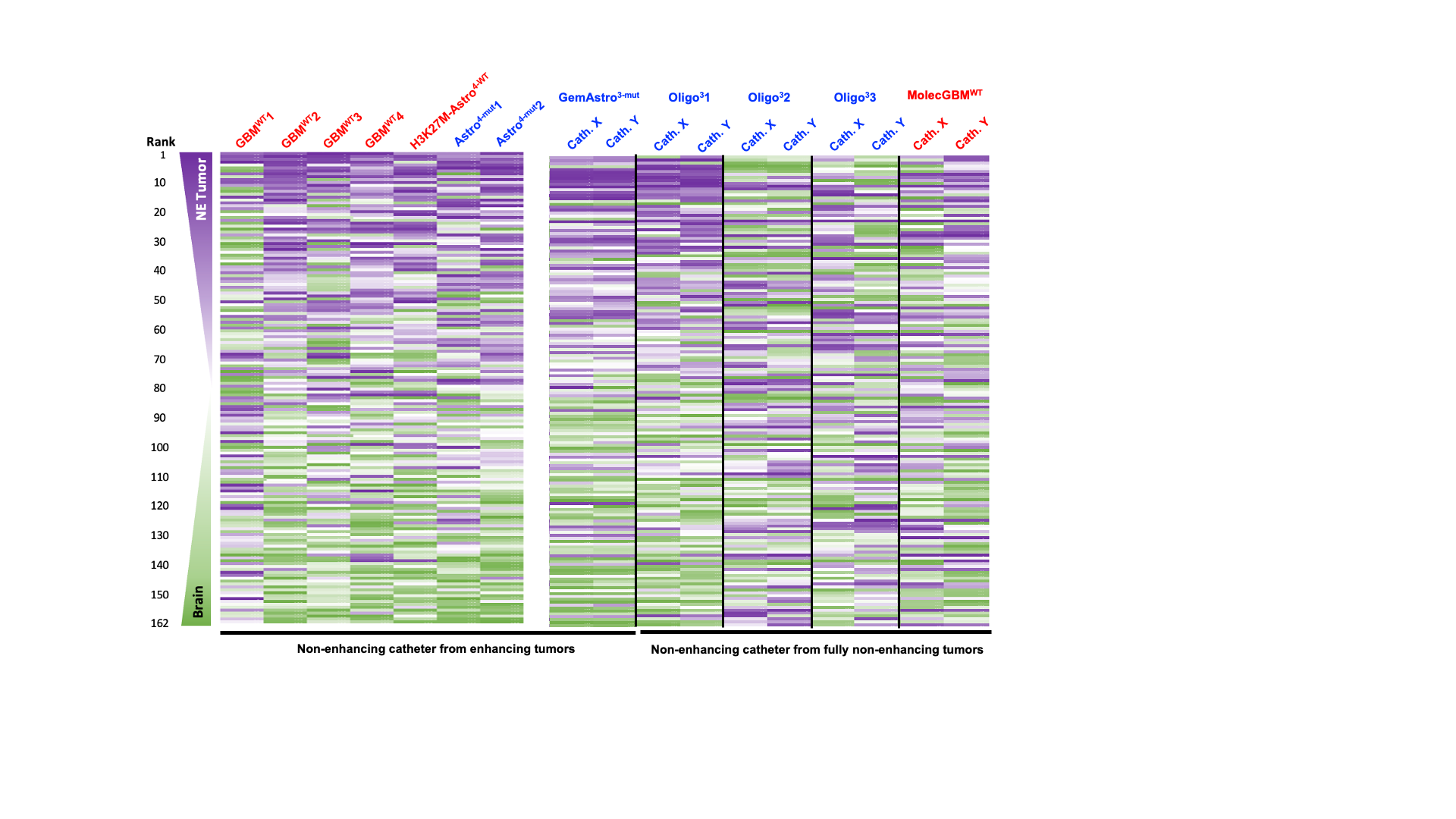

**Supplementary Figure S5. Non-enhancing catheter vs. brain metabolite signature – expanded cohort.**

The non-enhancing catheter versus brain metabolite signature from enhancing tumors (heatmap for 7 patients replicated from **Fig 3Ci** for clarity) was evaluated in an independent cohort of patients who only had non-enhancing tumor and brain microdialysate samples. Ranked metabolite lists were created for these five patients (right) between non-enhancing catheters (Cath X or Y) and brain (Z). The ranked order is conveyed as a heat map (1: non-enhancing, purple, to 162: brain, green) based on the ranked average of the depicted seven patients.

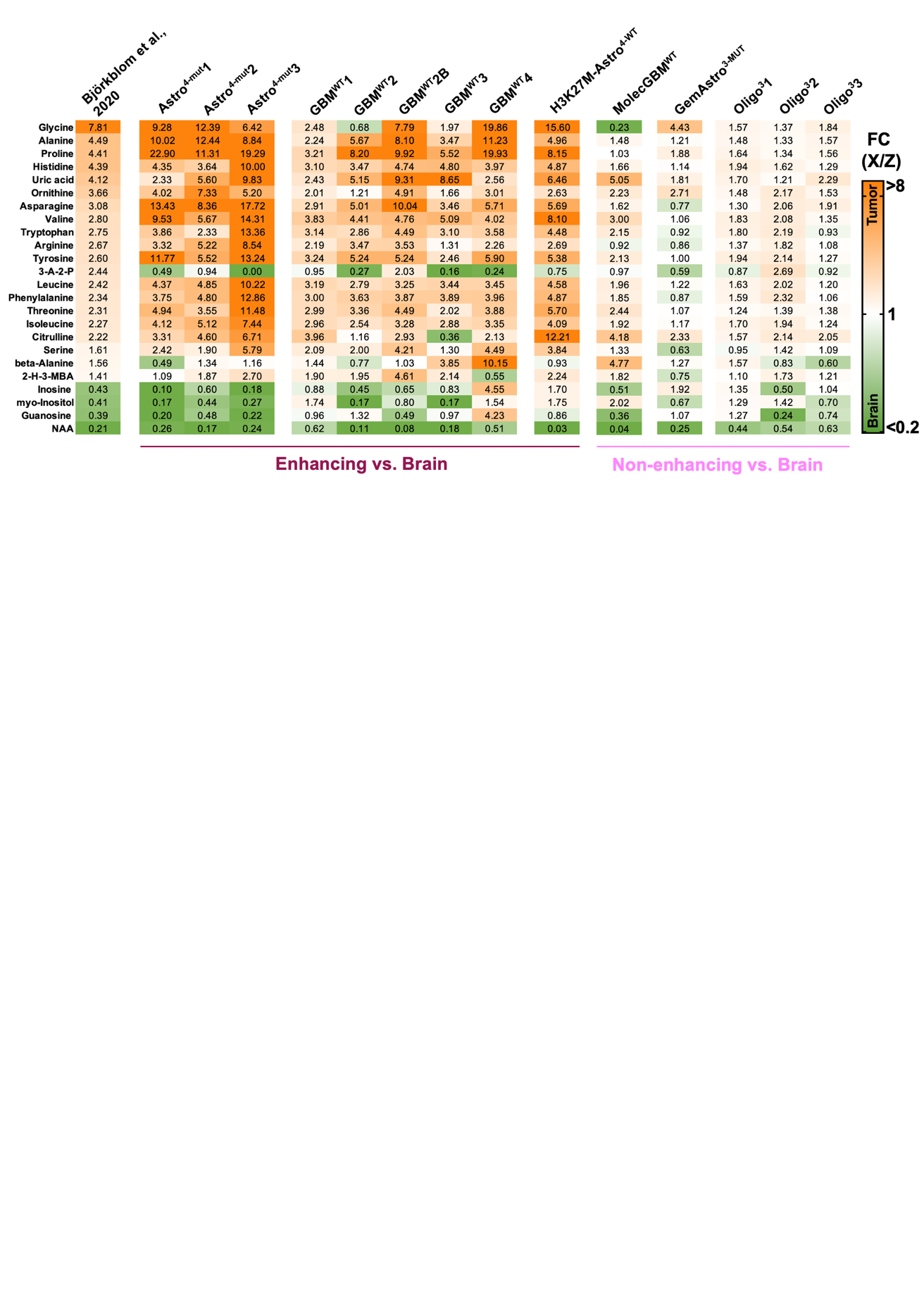

**Supplementary Figure S6. Tumor versus brain metabolites as reported by Björkblom et al., 2020**. The fold change (tumor versus brain) of significantly differentially abundant metabolites as reported by Björkblom et al, 2020 were compared to the fold changes observed between tumor and brain in our current data set for catheters X vs. Z. Fold change values are color coded from ≤ 0.2 (green = brain) to ≥ 8 (orange = tumor). 2-H-3-MBA: 2-hydroxy-3-methylbutyric acid; 3-A-2-P: 3-amino-2-piperidone; NAA: N-acetyl-L-aspartic acid.

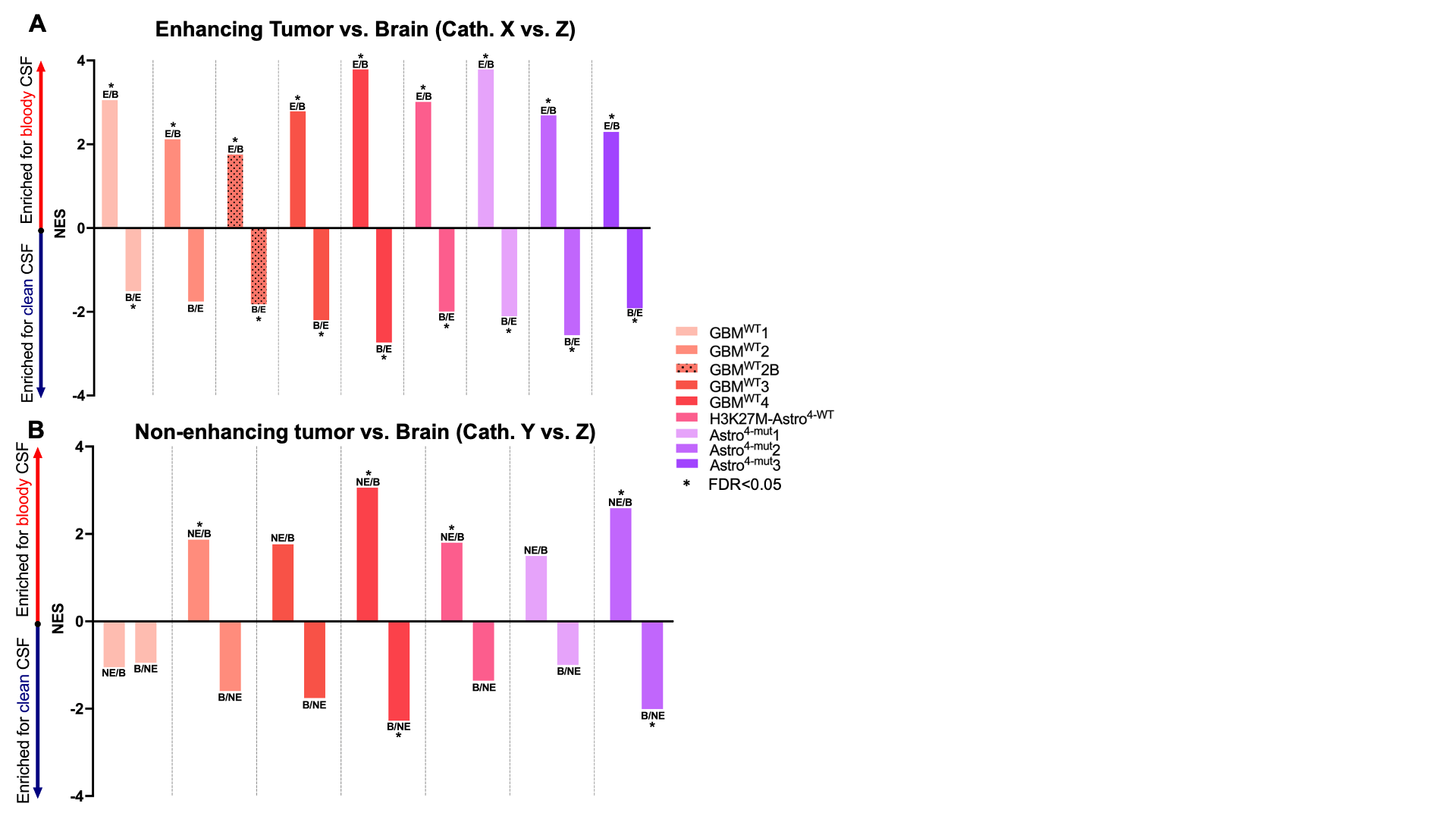

**Supplementary Figure S7. Plasma-derived metabolites in enhancing and non-enhancing versus brain microdialysates.**

Enrichment Analysis was utilized to determine the enrichment of each patient’s (A) enhancing tumor or (B) non-enhancing tumor versus brain (ranked list) for bloody versus clean CSF (metabolite set). Positive normalized enrichment scores (NES) indicate metabolic similarities to bloody CSF; negative enrichment scores indicate metabolic similarities to clean CSF (*= FDR<0.05).

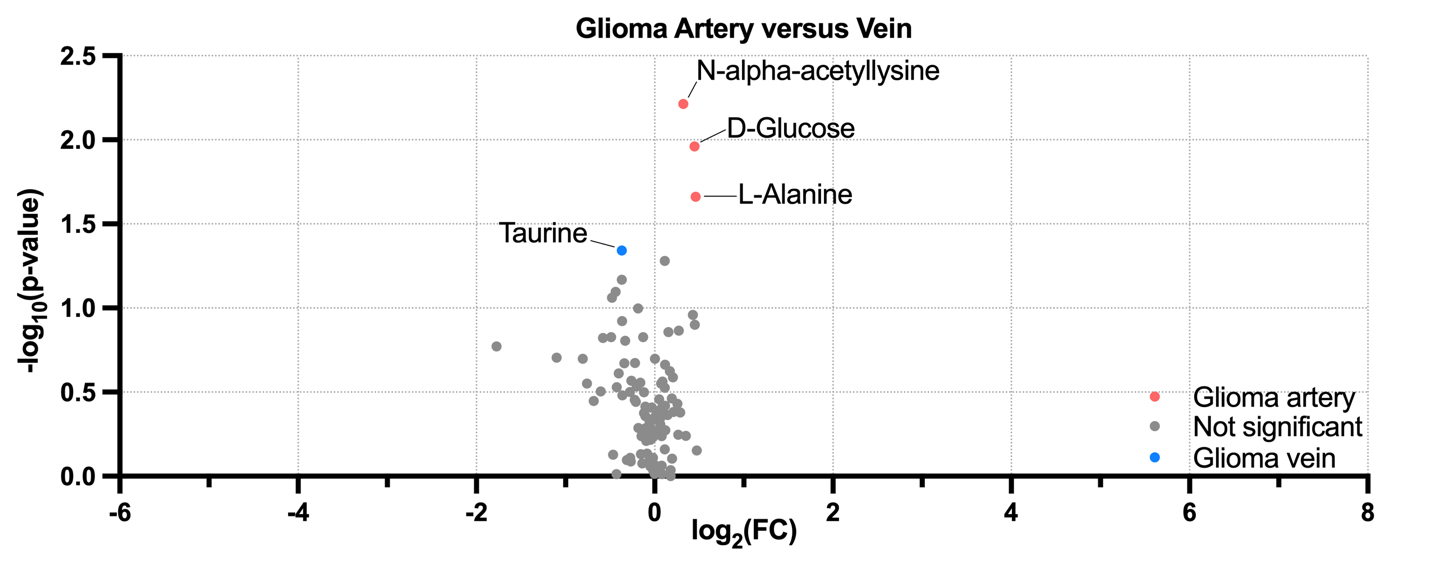

**Supplementary Figure S8: Analysis of publicly available data (Xiong, et al, 2020) comparing the venous and arterial glioma metabolome.** Paired t-tests and fold-changes were calculated between paired venous and arterial blood samples reported for 12 patients.

**Supplemental Table 1.** Intraoperative technical information for each patient.

| **Patient** | **Age Range/Sex** | **Tumor Location** | **Catheter Length** | **Microperfusate** |
| --- | --- | --- | --- | --- |
| Oligo^2^ | 20s/F | Left frontal | A&C: 10 mm; B: 20 mm | Lactated Ringer’s with 3% dextran 40 |
| Oligo^3^1 | 30s/F | Left frontal | A&B: 20 mm; C: 10 mm | Lactated Ringer’s with 3% dextran 40 |
| Oligo^3^2 | 40s/M | Right frontal | 10 mm | Artificial CSF with 3% Dextran 500 |
| Oligo^3^3 | 30s/M | Left frontal | 10 mm | 3% Albumin A in Plasmalyte A |
| GemAstro^3-mut^ | 60s/M | Left frontal | 10 mm | 3% Albumin A in Plasmalyte A |
| Astro^4-mut^1 | 40s/M | Right parietal | 10 mm | Lactated Ringer’s with 3% dextran 40 |
| Astro^4-mut^2 | 30s/F | Left frontoparietal | 10 mm | 3% Albumin A in Plasmalyte A |
| Astro^4-mut^3 | 30s/M | Right temporal | 10 mm | Artificial CSF with 3% Dextran 500 |
| H3K27M-Astro^4-WT^ | 40s/M | Right frontal | 10 mm | Artificial CSF with 3% Dextran 500 |
| GBM^WT^1 | 50s/M | Left temporal | 20 mm | Lactated Ringer’s with 3% dextran 40 |
| GBM^WT^2/2B | 40s/M | Left frontal | 10 mm | Artificial CSF with 3% Dextran 500 |
| GBM^WT^3 | 60s/F | Right temporal | 10 mm | Artificial CSF with 3% Dextran 500 |
| GBM^WT^4 | 40s/F | Right parietal | 10 mm | Artificial CSF with 3% Dextran 500 |
| MolecGBM^WT^ | 70s/M | Right insular/frontal | 10 mm | Artificial CSF with 3% Dextran 500 |

**Supplemental Table 2.** FDR values for enrichment analyses of enhancing tumor vs. brain ranked lists across patients (from Figure 3Aii).

|  | GBM^WT^1:  X vs. Z | GBM^WT^2:  X vs. Z | GBM^WT^2B:  X vs. Z | GBM^WT^3:  X vs. Z | GBM^WT^4:  X vs. Z | H3K27M-Astro^4-WT^:  X vs. Z | Astro^4-mut^1:  X vs. Z | Astro^4-mut^2:  X vs. Z | Astro^4-mut^3:  X vs. Z |
| --- | --- | --- | --- | --- | --- | --- | --- | --- | --- |
| GBM^WT^1: X vs. Z | 0.E+00 | 3.E-04 | 6.E-04 | 0.E+00 | 0.E+00 | 0.E+00 | 0.E+00 | 0.E+00 | 0.E+00 |
| GBM^WT^2: X vs. Z | 0.E+00 | 0.E+00 | 0.E+00 | 0.E+00 | 0.E+00 | 0.E+00 | 0.E+00 | 0.E+00 | 0.E+00 |
| GBM^WT^2: X vs. Z | 0.E+00 | 0.E+00 | 0.E+00 | 0.E+00 | 0.E+00 | 0.E+00 | 0.E+00 | 0.E+00 | 0.E+00 |
| GBM^WT^3: X vs. Z | 0.E+00 | 0.E+00 | 0.E+00 | 0.E+00 | 0.E+00 | 0.E+00 | 0.E+00 | 0.E+00 | 0.E+00 |
| GBM^WT^4: X vs. Z | 1.E-04 | 3.E-02 | 5.E-03 | 3.E-03 | 0.E+00 | 1.E-04 | 4.E-02 | 0.E+00 | 7.E-03 |
| H3K27M-Astro^4-WT^: X vs. Z | 0.E+00 | 0.E+00 | 0.E+00 | 0.E+00 | 0.E+00 | 0.E+00 | 0.E+00 | 0.E+00 | 0.E+00 |
| Astro^4-mut^1: X vs. Z | 0.E+00 | 0.E+00 | 0.E+00 | 0.E+00 | 0.E+00 | 0.E+00 | 0.E+00 | 0.E+00 | 0.E+00 |
| Astro^4-mut^2: X vs. Z | 0.E+00 | 0.E+00 | 0.E+00 | 0.E+00 | 0.E+00 | 0.E+00 | 0.E+00 | 0.E+00 | 0.E+00 |
| Astro^4-mut^3: X vs. Z | 0.E+00 | 0.E+00 | 0.E+00 | 0.E+00 | 0.E+00 | 0.E+00 | 0.E+00 | 0.E+00 | 0.E+00 |
| GBM^WT^1: Z vs. X | 0.E+00 | 0.E+00 | 0.E+00 | 0.E+00 | 8.E-04 | 0.E+00 | 0.E+00 | 0.E+00 | 0.E+00 |
| GBM^WT^2: Z vs. X | 0.E+00 | 0.E+00 | 0.E+00 | 0.E+00 | 2.E-03 | 0.E+00 | 0.E+00 | 0.E+00 | 0.E+00 |
| GBM^WT^2B: Z vs. X | 0.E+00 | 0.E+00 | 0.E+00 | 0.E+00 | 0.E+00 | 0.E+00 | 0.E+00 | 0.E+00 | 0.E+00 |
| GBM^WT^3: Z vs. X | 6.E-05 | 0.E+00 | 0.E+00 | 0.E+00 | 5.E-03 | 7.E-05 | 0.E+00 | 0.E+00 | 0.E+00 |
| GBM^WT^4: Z vs. X | 0.E+00 | 0.E+00 | 0.E+00 | 0.E+00 | 0.E+00 | 0.E+00 | 7.E-04 | 0.E+00 | 8.E-05 |
| H3K27M-Astro^4-WT^: Z vs. X | 0.E+00 | 0.E+00 | 0.E+00 | 0.E+00 | 0.E+00 | 0.E+00 | 0.E+00 | 0.E+00 | 0.E+00 |
| Astro^4-mut^1: Z vs. X | 0.E+00 | 0.E+00 | 0.E+00 | 0.E+00 | 4.E-02 | 0.E+00 | 0.E+00 | 0.E+00 | 0.E+00 |
| Astro^4-mut^2: Z vs. X | 0.E+00 | 0.E+00 | 0.E+00 | 0.E+00 | 0.E+00 | 0.E+00 | 0.E+00 | 0.E+00 | 0.E+00 |
| Astro^4-mut^3: Z vs. X | 0.E+00 | 0.E+00 | 0.E+00 | 0.E+00 | 3.E-02 | 0.E+00 | 0.E+00 | 0.E+00 | 0.E+00 |
| Tumor (Björkblom, 2020) | 1.E-03 | 0.E+00 | 0.E+00 | 4.E-04 | 2.E-03 | 2.E-03 | 0.E+00 | 0.E+00 | 0.E+00 |

**Supplemental Table 3.** FDR values for enrichment analyses of enhancing tumor vs. non-enhancing tumor ranked lists across patients (from Figure 3Bii).

|  | GBM^WT^1:  X vs. Y | GBM^WT^2:  X vs. Y | GBM^WT^3:  X vs. Y | GBM^WT^4:  X vs. Y | H3K27M-Astro^4-WT^:  X vs. Y | Astro^4-mut^1:  X vs. Y | Astro^4-mut^2:  X vs. Y |
| --- | --- | --- | --- | --- | --- | --- | --- |
| GBM^WT^1: X vs. Y | 0.E+00 | 1.E-02 | 9.E-04 | 0.E+00 | 3.E-04 | 8.E-01 | 2.E-04 |
| GBM^WT^2: X vs. Y | 5.E-03 | 0.E+00 | 0.E+00 | 0.E+00 | 0.E+00 | 4.E-04 | 9.E-04 |
| GBM^WT^3: X vs. Y | 0.E+00 | 0.E+00 | 0.E+00 | 0.E+00 | 0.E+00 | 0.E+00 | 0.E+00 |
| GBM^WT^4: X vs. Y | 0.E+00 | 0.E+00 | 0.E+00 | 0.E+00 | 0.E+00 | 3.E-04 | 0.E+00 |
| H3K27M-Astro^4-WT^: X vs. Y | 4.E-04 | 6.E-04 | 0.E+00 | 0.E+00 | 0.E+00 | 0.E+00 | 0.E+00 |
| Astro^4-mut^1: X vs. Y | 9.E-01 | 7.E-04 | 3.E-04 | 2.E-04 | 6.E-05 | 0.E+00 | 2.E-04 |
| Astro^4-mut2^: X vs. Y | 0.E+00 | 0.E+00 | 0.E+00 | 0.E+00 | 0.E+00 | 0.E+00 | 0.E+00 |
| GBM^WT^1: Y vs. X | 0.E+00 | 9.E-02 | 3.E-02 | 2.E-02 | 1.E-03 | 1.E-01 | 3.E-03 |
| GBM^WT^2: Y vs. X | 5.E-01 | 0.E+00 | 0.E+00 | 4.E-04 | 0.E+00 | 0.E+00 | 2.E-03 |
| GBM^WT^3: Y vs. X | 3.E-03 | 0.E+00 | 0.E+00 | 0.E+00 | 0.E+00 | 0.E+00 | 0.E+00 |
| GBM^WT^4: Y vs. X | 3.E-03 | 0.E+00 | 0.E+00 | 0.E+00 | 0.E+00 | 3.E-01 | 2.E-03 |
| H3K27M-Astro^4-WT^: Y vs. X | 1.E-01 | 0.E+00 | 0.E+00 | 0.E+00 | 0.E+00 | 0.E+00 | 7.E-05 |
| Astro^4-mut^1: Y vs. X | 2.E-01 | 4.E-04 | 3.E-04 | 5.E-01 | 8.E-03 | 0.E+00 | 0.E+00 |
| Astro^4-mut2^: Y vs. X | 4.E-01 | 4.E-04 | 0.E+00 | 0.E+00 | 0.E+00 | 0.E+00 | 0.E+00 |

**Supplemental Table 4.** FDR values for enrichment analyses of Non-enhancing tumor vs. brain ranked lists across patients (from Figure 3cii).

|  | GBM^WT^1:  Y vs. Z | GBM^WT^2:  Y vs. Z | GBM^WT^3:  Y vs. Z | GBM^WT^4:  Y vs. Z | H3K27M-Astro^4-WT^:  Y vs. Z | Astro^4-mut^1:  Y vs. Z | Astro^4-mut^2:  Y vs. Z |
| --- | --- | --- | --- | --- | --- | --- | --- |
| GBM^WT^1: Y vs. Z | 0.E+00 | 4.E-01 | 1.E-01 | 1.E-01 | 6.E-02 | 7.E-01 | 4.E-01 |
| GBM^WT^2: Y vs. Z | 2.E-01 | 0.E+00 | 1.E-01 | 5.E-03 | 0.E+00 | 2.E-03 | 2.E-04 |
| GBM^WT^3: Y vs. Z | 7.E-02 | 6.E-04 | 0.E+00 | 0.E+00 | 3.E-02 | 6.E-01 | 8.E-02 |
| GBM^WT^4: Y vs. Z | 1.E-01 | 8.E-03 | 0.E+00 | 0.E+00 | 0.E+00 | 3.E-02 | 2.E-01 |
| H3K27M-Astro^4-WT^: Y vs. Z | 1.E-02 | 0.E+00 | 3.E-01 | 7.E-04 | 0.E+00 | 3.E-01 | 4.E-01 |
| Astro^4-mut^1: Y vs. Z | 2.E-01 | 1.E-02 | 8.E-02 | 5.E-01 | 2.E-02 | 0.E+00 | 3.E-04 |
| Astro^4-mut2^: Y vs. Z | 4.E-01 | 0.E+00 | 6.E-01 | 7.E-03 | 1.E-01 | 0.E+00 | 0.E+00 |
| GBM^WT^1: Z vs. Y | 0.E+00 | 4.E-02 | 9.E-02 | 2.E-02 | 0.E+00 | 2.E-01 | 2.E-01 |
| GBM^WT^2: Z vs. Y | 6.E-01 | 0.E+00 | 1.E+00 | 3.E-01 | 7.E-02 | 0.E+00 | 9.E-03 |
| GBM^WT^3: Z vs. Y | 5.E-01 | 7.E-01 | 0.E+00 | 2.E-01 | 3.E-01 | 2.E-02 | 6.E-01 |
| GBM^WT^4: Z vs. Y | 6.E-02 | 0.E+00 | 8.E-02 | 0.E+00 | 0.E+00 | 3.E-01 | 2.E-02 |
| H3K27M-Astro^4-WT^: Z vs. Y | 6.E-03 | 4.E-02 | 7.E-01 | 1.E-04 | 0.E+00 | 8.E-01 | 2.E-01 |
| Astro^4-mut^1: Z vs. Y | 2.E-01 | 5.E-04 | 7.E-01 | 4.E-01 | 7.E-01 | 0.E+00 | 6.E-04 |
| Astro^4-mut2^: Z vs. Y | 8.E-01 | 1.E-02 | 2.E-01 | 8.E-02 | 3.E-01 | 1.E-04 | 0.E+00 |
